# Short-range airborne route dominates exposure of respiratory infection during close contact

**DOI:** 10.1101/2020.03.16.20037291

**Authors:** Wenzhao Chen, Nan Zhang, Jianjian Wei, Hui-Ling Yen, Yuguo Li

## Abstract

A susceptible person experiences the highest exposure risk of respiratory infection when he or she is in close proximity with an infected person. The large droplet route has been commonly believed to be dominant for most respiratory infections since the early 20^th^ century, and the associated droplet precaution is widely known and practiced in hospitals and in the community. The mechanism of exposure to droplets expired at close contact, however, remains surprisingly unexplored. In this study, the exposure to exhaled droplets during close contact (< 2 m) via both the short-range airborne and large droplet sub-routes is studied using a simple mathematical model of expired flows and droplet dispersion/deposition/inhalation, which enables the calculation of exposure due to both deposition and inhalation. The short-range airborne route is found to dominate at most distances studied during both talking and coughing. The large droplet route only dominates when the droplets are larger than 100 μm and when the subjects are within 0.2 m while talking or 0.5 m while coughing. The smaller the exhaled droplets, the more important the short-range airborne route. The large droplet route contributes less than 10% of exposure when the droplets are smaller than 50 μm and when the subjects are more than 0.3 m apart, even while coughing.

**Practical implications:** Our simple but novel analysis shows that conventional surgical masks are not effective if most infectious viruses are contained in fine droplets, and non-conventional intervention methods such as personalised ventilation should be considered as infection prevention strategies given the possible dominance of the short-range airborne route, although further clinical evidence is needed.

**Nomenclature:** *Subscript:* *i* Droplets of different diameter groups (*i* = 1, 2, …, N) *LD* Large droplet route *SR* Short-range airborne route

*Symbols:* *A*_0_ Area of source mouth [m^2^] *AE* Aspiration efficiency [-] *Ar*_0_ Archimedes number [-] *b*_*g*_ Gaussian half width [m] *b*_*t*_ Top-hat half width [m] *C*_*D*_ Drag coefficient [-] *C*_*I*_ Specific heat of liquid [J•kg^-1^•K^-1^] *C*_*s*_ Specific heat of solid [J•kg^-1^•K^-1^] *C*_*T*_ Correction factor for diffusion coefficient due to temperature dependence [-] *d*_*d*_ Droplet diameter [m] *d*_*d*0_ Droplet initial diameter [m] *d*_*e*1_ Major axis of eye ellipse [m] *d*_*e*2_ Minor axis of eye ellipse [m] *d*_h_ Characteristic diameter of human head [m] *d*_*m*_ Mouth diameter [m] *d*_*n*_ Nostril diameter [m] *D*_∞_ Binary diffusion coefficient far from droplet [m^2^•s^-1^] *DE* Deposition efficiency [-] *e*_*LD*_ Exposure due to large droplet route [μL] *e*_*SR*_ Exposure due to short-range airborne route [μL] *g* Gravitational acceleration [m•s^-2^] *I*_*v*_ Mass current [kg•s^-1^] *IF* Inhalation fraction [-] *K*_*c*_ Constant (=0.3) [-] *K*_*g*_ Thermal conductivity of air [W•m^-1^•K^-1^] *LS* Exposure ratio between large droplet and short-range airborne [-] *L*_*v*_ Latent heat of vaporization [J•kg^-1^] *m*_*d*_ Droplet mass [kg] *m*_*I*_ Mass of liquid in a droplet [kg] *m*_*s*_ Mass of solid in a droplet [kg] *M*_0_ Jet initial momentum [m^4^•s^-2^] *M*_*W*_ Molecular weight of H_2_O [kg•mol^-1^] *MF* Membrane fraction [-] *n* Number of droplets [n] *n*_0_ Number of droplets expelled immediately at mouth [n] *N*_*in*_ Number of droplets entering the inhalation zone [n] *N*_*m*_ Number of droplets potentially deposited on mucous membranes [n] *N*_*t*_ Total number of released droplets [n] Nu Nusselt number [-] *p* Total pressure [Pa] *p*_*v*∞_ Vapour pressure distant from droplet surface [Pa] *p*_*vs*_ Vapour pressure at droplet surface [Pa] *Q*_*jet*_ Jet flow rate [m^3^•s^-1^] *r* Radial distance away from jet centreline [m] *r*_*d*_ Droplet radius [m] *R* Radius of jet potential core [m] *R*_g_ Universal gas constant [J•K^-1^•mol^-1^] *s* Jet centreline trajectory length [m] *S*_*in*_ Width of region on sampler enclosed by limiting stream surface [m] Sh Sherwood number [-] *St*_*c*_ Stokes number in convergent part of air stream [-] *St*_h_ Stokes number for head [-] *St*_*m*_ Stokes number for mouth [-] *t* Time [s] *T*_0_ Initial temperature of jet [K] *T*_∞_ Ambient temperature [K] *T*_*d*_ Droplet temperature [K] *u*_0_ Initial velocity at mouth outlet [m•s^-1^] *u*_*d*_ Droplet velocity [m•s^-1^] *u*_*g*_ Gaussian velocity [m•s^-1^] *u*_*g*as_ Gas velocity [m•s^-1^] *u*_*gc*_ Gaussian centreline velocity [m•s^-1^] *u*_*in*_ Inhalation velocity [m•s^-1^] *u*_*t*_ Top-hat velocity [m•s^-1^] *v*_*p*_ Individual droplet volume considering evaporation [m^3^] *x* Horizontal distance between source and target [m] *z* Jet vertical centreline position [m] *ρ*_0_ Jet initial density [kg•m^-3^] *ρ*_∞_ Ambient air density [kg•m^-3^] *ρ*_*d*_ Droplet density [kg•m^-3^] *ρ*_*g*_ Gas density [kg•m^-3^] *Δρ* Density difference between jet and ambient air [kg•m^-3^] *μ*_*g*_ Gas dynamic viscosity [Pa•s] φ Sampling ratio in axisymmetric flow system [-] *α*_*c*_ Impaction efficiency in convergent part of air stream [-]

## 1. Introduction

Despite significant progress in medicine and personal hygiene, seasonal respiratory infections such as influenza remain a significant threat to human health as a result of more frequent social contact and rapid genetic evolution of microbes. Disease transmission is a complex and interdisciplinary process related to microbiology, environmental and social science. The respiratory activities of an infected person (infected), such as talking and coughing, release expiratory droplets that contain infectious pathogens, and these expired droplets can be the medium for transmitting infection. Exposure to these droplets leads to risk of infection and/or disease. Three possible routes of transmission have been widely recognised and studied: the airborne, fomite and large droplet (or droplet-borne) routes [1]. The former two are examples of distant infection, whilst the latter occurs with close contact.

When a susceptible individual is in close contact with an infected, the risk of exposure to exhaled droplets is expected to be at its greatest. The concentration of exhaled droplets is higher in expired jets than in ambient air. Brankston et al. [2] suggested that transmission of influenza is most likely to occur at close contact. Close interpersonal contact is ubiquitous in daily life, such as in offices [3], schools and homes. Although it varies between cultures [4], the interpersonal distance is normally within 1.5-2 m. Close contact in itself is not a transmission route, but a facilitating event for droplet transmission. Note that the use of “droplets” in the remaining text refers to all sizes, down to and including all fine droplets, such as the sub-micron ones. Two major sub-routes are possible in close contact transmission. The large droplet sub-route refers to the deposition of large droplets on the lip/eye/nostril mucosa of another person at close proximity, resulting in his or her self-inoculation. Dry surroundings enable the exhaled droplets to evaporate, and some rapidly shrink to droplet nuclei. The fine droplets and droplet nuclei can also be directly inhaled, which is the short-range airborne sub-route. Both sub-routes involve direct exposure to the expired jet, which is affected by the interacting exhalation/inhalation flows of the two persons. For example, head movement can change the orientation of the expired flow, and the mode of breathing affects the interaction. The significance of breathing mode (mouth/nose) and distance between people in cross-infection risk has been widely studied [5]. Body thermal plumes can also interact with the expired jet from the infected and with potential inhalation of the flow by the susceptible person [1].

### It remains an open question whether either of the two sub-routes is dominant, or both are important

The large droplet route has been believed to be dominant for most respiratory infections [2] since Flügge [6] and Chapin [7]. Some epidemiological studies have even assumed respiratory infections to be due to large droplets whenever close contact transmission is observed [8]. Liu et al. [9] showed that both the large droplet route and the short-range airborne route can be important within 1.5 m. However, their computational fluid dynamics (CFD) modelling considered only a very small number of droplets, and the frequency of droplet deposition on the mucosa was not estimated. Except for that study by Liu et al. [9], comparison of the two sub-routes has rarely been reported. In the general discipline of exposure science, particle inhalability has been studied in depth, due to the potential health impact of particles when inhaled; see Vincent [10] for a comprehensive review. There are also considerable data on particle inhalability in humans. However, the short-range airborne route, or expired droplet inhalability at close contact, that we consider here differs from conventional particle inhalability (e.g., [11]) in at least two aspects. First, it is not the room air flow that affects inhalability, but the expired air stream from the source person. The inhalability depends upon whether the susceptible person’s mouth or nose is located within or partially within the cone of the expired jet from the source person. The size of the expired droplets changes due to evaporation after being exhaled and before being inhaled or deposited on the mucous membranes. Large droplet deposition on mucous membranes has rarely been studied in combination with their inhalation. Kim et al. [12] investigated aerosol-based drug delivery for a 7-month infant, taking both large droplet and short-range routes into account using CFD. They found that droplet deposition was determined more by head direction than by inhalation, suggesting the importance of close contact parameters.

The importance of identifying the dominant/important sub-route(s) in close contact is obvious. There are significant implications for the choice and development of effective intervention measures. If the short-range airborne sub-route is dominant, a face mask (a typical droplet precaution) will not be sufficient because these masks cannot remove fine droplets. This study aims to tackle the question of the relative importance of the two exposure sub-routes using simple analysis.

## 2. Methods

A mathematical model is developed here, based on the simple dynamics of expired jets. As in inhalability studies, we consider the droplet inhalation and deposition processes as particle sampling (e.g. [13]).

The large droplet route and short-range airborne route are illustrated in Figure 1 for two standing persons, who might be in conversation or simply in face-to-face contact, within less than 2 m. One individual is identified as the source (the infected) and the other as the target (the susceptible person). Droplets can be directly deposited on the susceptible person’s facial membranes (eyes, nostrils and mouth; i.e., the large droplet sub-route), whilst those inhaled via oral breathing are categorised into the short-range airborne sub-route. The terminologies “large droplet” and “short-range airborne” here apply to an overall droplet size range, and each size of droplets (as shown in Figure 2) will have opportunities to be deposited or inhaled, regardless of its diameter. Note that these two sub-routes are considered as two separate processes; that is, the large droplet and short-range airborne routes do not happen simultaneously, and infection occurs through the mouth in both cases. The environmental conditions include air temperature (25°C), relative humidity (RH = 50%) and atmospheric pressure (101,325 Pa). The room air flows are also not considered (i.e., background air at 0 m/s). Droplets were released from a height of 1.75 m, considering that both individuals were standing.

**Figure 1.**
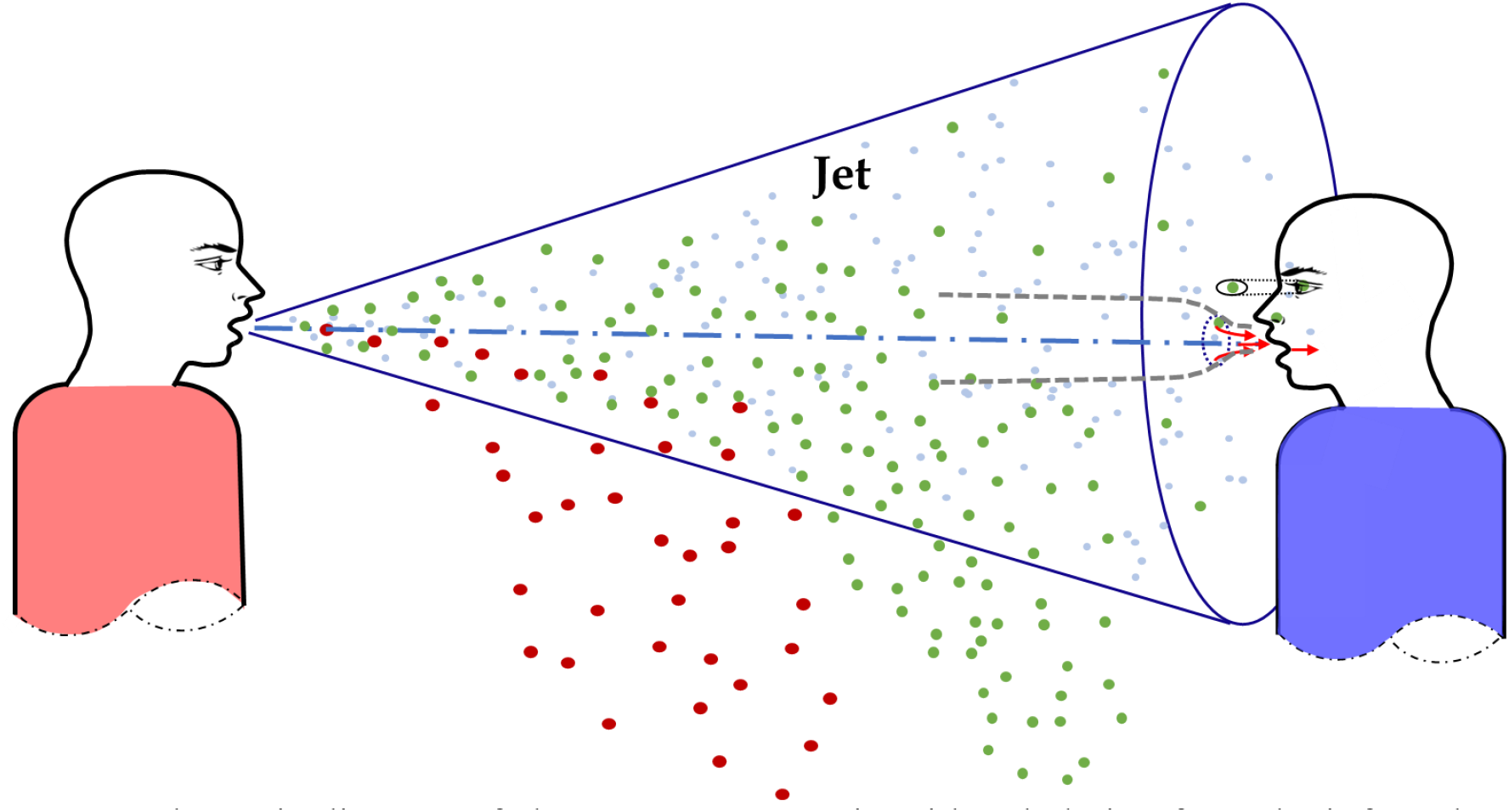
Schematic diagram of close contact scenario with exhalation from the infected (left) and inhalation through the mouth of the susceptible person (right).

**Figure 2.**
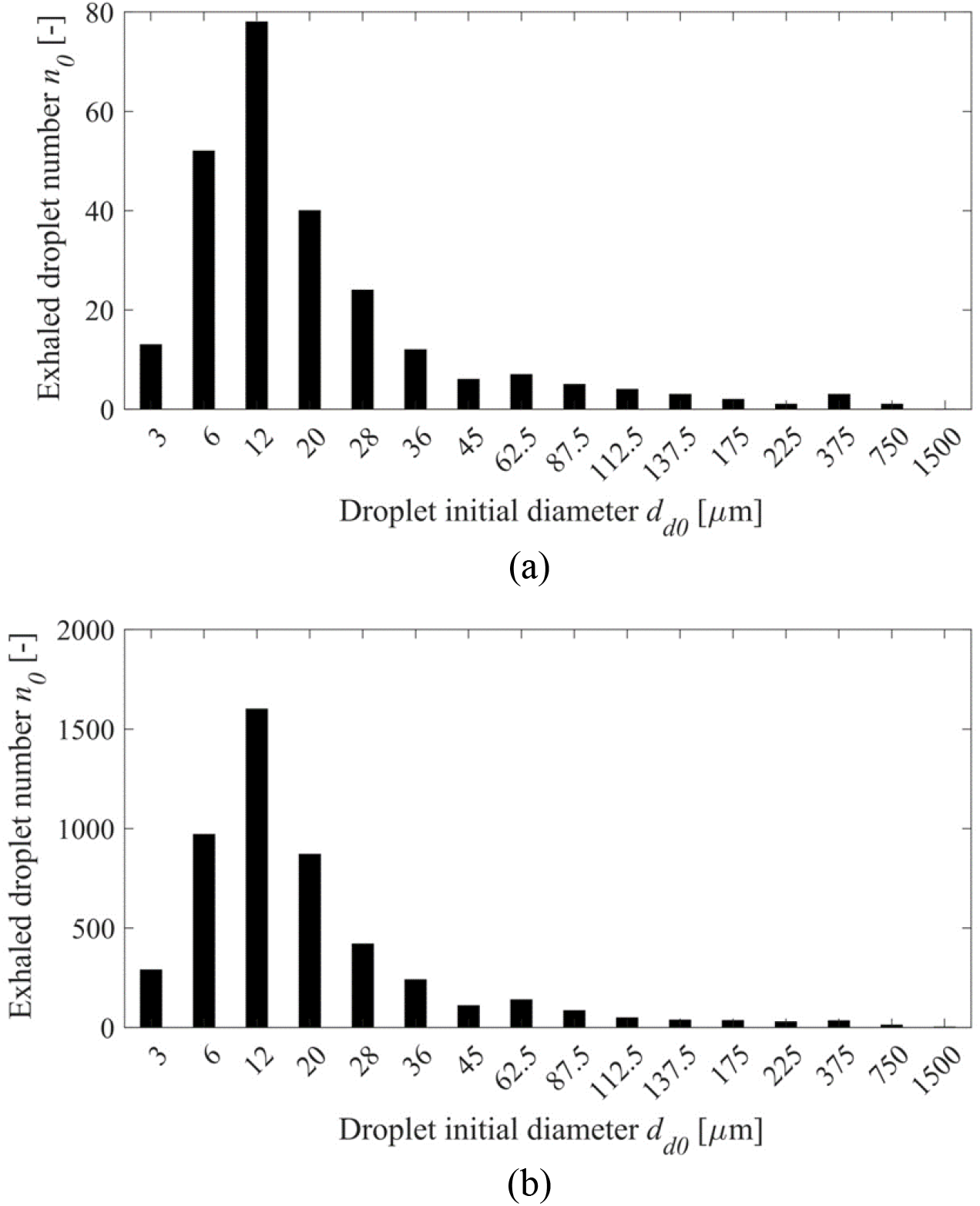
Number distributions of exhaled droplets at the point of mouth opening. (a) Talking (counting from ‘1’ to ‘100’ once) [n]; (b) Coughing once [n].

The exposure is defined as the total volume of droplets to which the susceptible person is exposed, in units of μL. The riskiest situation was investigated here, that in which the susceptible person is in *direct* face-to-face contact with the source. For the short-range airborne route, we assumed that the target took a breath exactly when the droplet-laden air flow exhaled by the infected reached him or her; for the large droplet route, the susceptible person was assumed to hold his or her breath with the mouth open. The two mouths are at the same height; see Figure 1. Hence, we studied perhaps the worst scenario in terms of large droplet transmission. Our model considers the spread of the exhalation jet, and the dispersion and evaporation of expired droplets, as an example of aerosol sampling, a process analogous to inhalation and consistent with human facial features. We used Matlab for implementing the prediction. The used models in terms of airflow and particle deposition have been previously validated.

### 2.1 Exposure calculation

The exposure via the large droplet and short-range airborne sub-routes at any horizontal distance *x* can be calculated as:

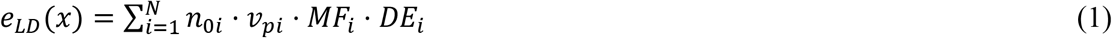

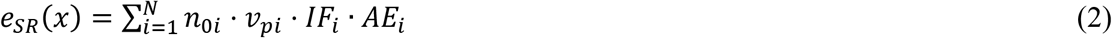

where subscript *LD* and *SR* denote the large droplet route and short-range airborne route, respectively; *i* stands for droplets sorted into groups based on diameter (*i* = 1, 2, …, N); *n*_0_ is the number of droplets expelled from the source mouth at the moment of exhalation; *v*_*p*_ is the individual droplet volume, taking into account evaporation; *MF* is the membrane fraction; *DE* is the deposition efficiency; *IF* is the inhalation fraction; and *AE* is the aspiration efficiency. These variables will be defined more specifically in the following sections. Our adopted index *IF* is not to be confused with intake fraction as used for example in Berlanga et al. [14]. The droplet number generated in expiratory activities has been measured by many researchers, e.g. [15-16]. To encompass a wide size range, the classical experimental dataset by Duguid [17] was adopted. The number distributions of different-sized droplets, as generated by two different exhalatory processes – counting out loud from ‘1’ to ‘100’ once (i.e., talking), and coughing once [17] – are shown in Figure 2 and refer to the *n*_0_ values. The total volumes of droplets released by talking and coughing are 0.32 μL and 7.55 μL respectively, which are calculated as the sum of droplet volume of each size. The diameters of the expired droplets may extend down to the submicron scale; however, we do not have access to a full and consistent set of data that include these submicron sizes.

To compare the relative contribution of the two sub-routes, an *LS* exposure ratio is defined at each horizontal distance *x*. If the *LS* ratio is greater than 1, the large droplet route dominates, and vice versa.

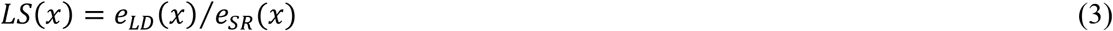

### 2.2 Velocity profiles in the expired jet

As a first approximation, the exhaled air flow from the infected source may be treated as a turbulent round jet, including a flow establishment zone and an established flow zone. The velocity profiles and the flow rate can be obtained by various jet theories. Given the fact that human exhalation can be complicated in terms of airflow fluctuations, individual differences, and exhaled flow directions [18], here we chose the classic jet formulas in Lee and Chu [19]. Let *s* be the centreline distance travelled by the jet and *d*_*m*_ the source mouth diameter (i.e. the jet opening, assumed to be 2 cm [20]). The maximum length of the flow establishment zone is 6.2*d*_*m*_.

In the flow establishment zone (*s* ≤ 6.2*d*_*m*_, Gaussian profile),

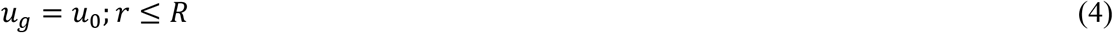

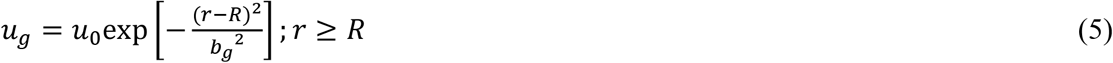

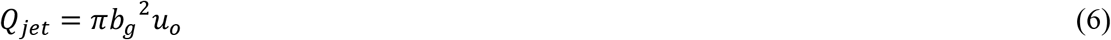

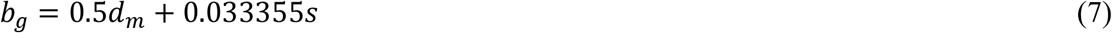

In the established flow zone (*s* > 6.2*d*_*m*_, Gaussian profile),

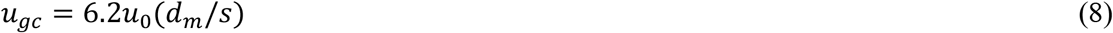

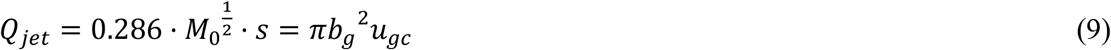

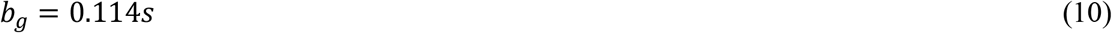

where *u*_*g*_ is the Gaussian velocity; *u*_0_ is the initial velocity at the source mouth outlet; *r* is the radial distance away from the jet centreline; *R* is the radius of the jet’s potential core; *b*_*g*_ is the Gaussian half width; *Q*_*jet*_ is the jet flow rate; *u*_*gc*_ is the Gaussian centreline velocity; and 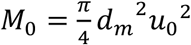 is the initial momentum. The velocities in the jet cone are used to calculate the trajectories of the expired droplets.

We also take the average velocity on a cross-section plane, which gives a top-hat profile. The average velocities are used to calculate the particle deposition.

In the flow establishment zone (*s* ≤ 6.2*d*_*m*_, top-hat profile),

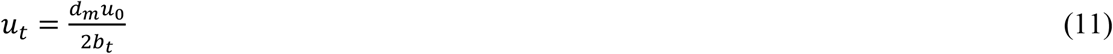

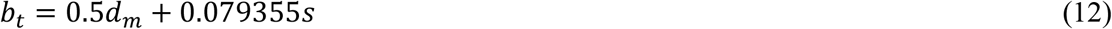

In the established flow zone (*s* > 6.2*d*_*m*_, top-hat profile),

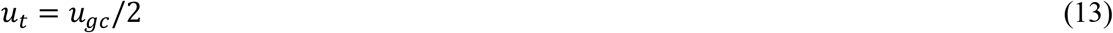

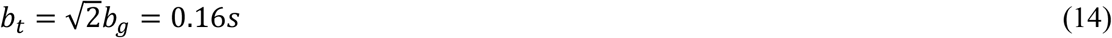

where *u*_*t*_ is the top-hat velocity; *b*_*t*_ is the top-hat half width.

We use the measured velocity of particles exhaled by different respiratory activities at the moment of mouth opening as reported by Chao et al. [21]. The average velocity at the mouth is 3.9 m/s for speaking and 11.7 m/s for coughing.

Under isothermal conditions, the jet centreline is assumed to be straight. The exhaled air temperature (assumed to be 35.1°C, averaged between patients with asthma and control subjects [22]) generally differs from the environmental temperature (typical room temperature 25°C). In this case, the jet trajectory would curve upwards [23] as in the following equations:

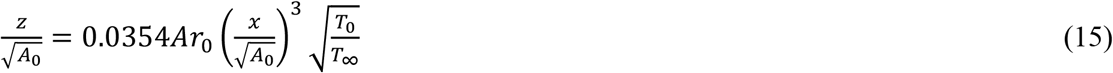

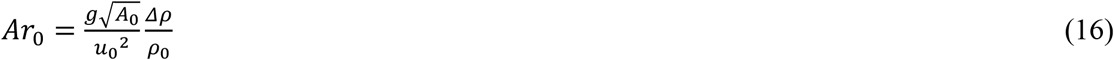

where *z* is the vertical centreline position; 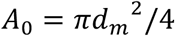 is the area of the source mouth; *Ar*_0_ is the Archimedes number; *T*_0_ is the initial temperature of the jet; *T*_∞_ is the ambient temperature; *g* is the gravitational acceleration; *ρ*_0_ is the jet initial density; *Δρ* = *ρ*_∞_ - *ρ*_0_ is the density difference between the jet and ambient air. Note that *x* is the horizontal distance between the source and the target, whilst *s* is the jet centreline trajectory length. Each *x* corresponds to an *s* value, and *s* is slightly larger than *x*.

### 2.3 Droplet evaporation and dispersion

To ensure a significant number of droplets depositing on face/membranes or entering the inhalation zone in calculating *MF* and *IF* (especially for droplets with large sizes), a total of 5000 droplets greater than 50 μm and 1600 droplets smaller than 50 μm were released. The simulation of droplet motion and evaporation was based on an existing model developed and validated in Wei and Li [20]. The governing equations for motion, mass flux and heat transfer are listed below. Droplets were modelled to be released randomly from the source mouth, which was divided into 1600 segments. The maximum distance studied is 2 m. Our prediction of droplet dispersion starts from the release at the source mouth and ends when falling on the ground or reaching 2 m. At each 0.1 m, data such as droplet velocity, position and size change were recorded.

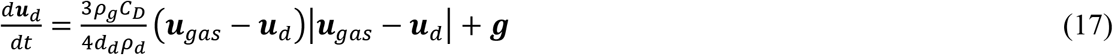

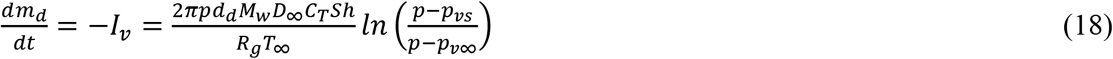

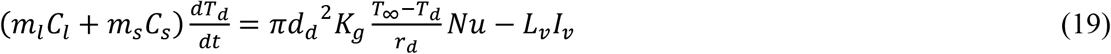

### 2.4 Deposition

The droplet membrane fraction *(MF*) is defined as the ratio of the number of droplets that are potentially deposited on the mucous membranes, *N*_*m*_, to the total number of released droplets, *N*_*t*_.

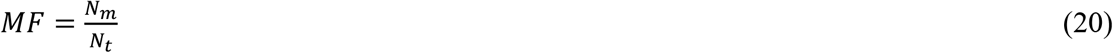

The process of deposition due to the large droplet route is illustrated in Figure 3b. The total surface area of the two eyes is 6 cm^2^ and that of the two nostrils is 2 cm^2^ [24]. The mouth is approximated as a circle with a diameter of 2 cm [20]. The total surface area of the eyes, nostrils and lips is approximately only 15 cm^2^ [25], compared with the average area for a head of 1300 cm^2^ [26]. A diagram of extracted facial features is shown in Figure 3a, with the eyes being treated as ellipses, the nose and mouth being circles. The vertical distance between the eyes and nose is 3.07 cm, and the distance between the eyes and mouth is 5.64 cm [27].

**Figure 3.**
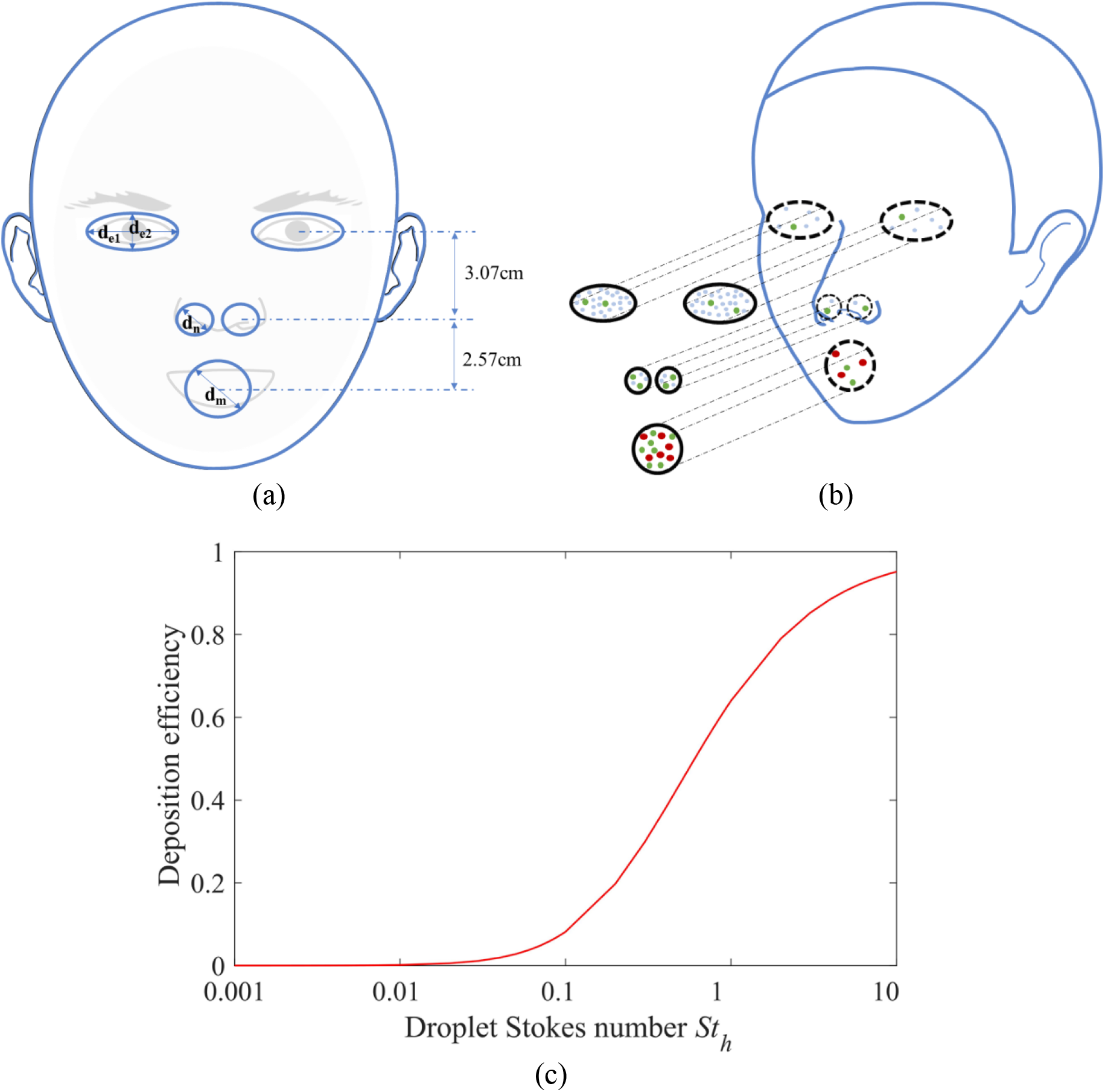
(a) Extraction of human facial features and their dimensions in our model (*d*_*e*1_ = 2.76 cm, *d*_*e*2_ = 1.38 cm, *d*_*n*_ = 1.13 cm, *d*_*m*_ = 2.00 cm); (b) illustration of the large droplet route, where only droplets deposited on mucous membranes are considered to result in exposure; note that only a fraction of droplets entering cylindrical volumes would eventually deposit; (c) variation of capture efficiency on a sphere with the Stokes number [28-31].

The number of droplets that are potentially deposited on the mucous membranes, *N*_*m*_, can be obtained by deciding whether a particular droplet is within the projected cylindrical volumes just in front of the eye ellipses or nose/mouth circles (see Figure 3b). Only a fraction of these droplets will deposit, while others would follow the airflow trajectory around the face. This enables the dispersion of droplets in the exhaled jet to be fully considered before arriving at the head of the susceptible person. This simple model does not consider the opening and closing of the eyes and mouth or that the nostril openings may not always be facing forward. By assuming that the eyes and mouth are always open and that droplets can always be directly deposited onto the nostrils, the model may overestimate the rate of large droplet deposition.

The deposition efficiency (*DE*) represents the probability of deposition, which is a function of the droplet Stokes number, a dimensionless number characterising the behaviour of droplets suspended in a fluid flow. Droplets with a small Stokes number follow the surrounding fluid flow, whilst those with a large Stokes number tend to continue their trajectory under inertia and are deposited. We approximate the head as a sphere. The droplet deposition efficiency on a sphere was first considered by Langmuir and Blodgett [28] (see Figure 3c). The model given by Equation (21) was in reasonable agreement with the experimental data of Walton and Woolcock [29]. The theory was further confirmed by measurement by Hähner et al. [30] and Waldenmaier [31]. The horizontal location differences among eyes, nostrils and mouth on the sphere were neglected. They were assumed to be on the same plane, although a spherical model was used in calculating the deposition.

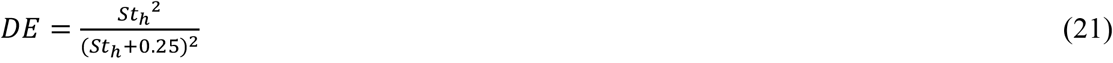

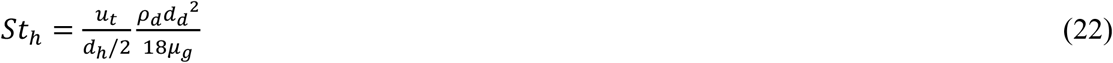

where *St*_h_ is the Stokes number for an approximate spherical head; *ρ*_*d*_ is the droplet density; *d*_*d*_ is the droplet diameter; *d*_h_ = 0.2 *m* is the characteristic diameter of the human head; *μ*_*g*_ is the gas dynamic viscosity. Considering the distributions of facial organs in the expired jet, *u*_*t*_ was used for Stokes number calculation.

### 2.5 Inhalation

The inhalation process is treated as an anisokinetic sampling process, with the human head approximated as a spherical aerosol sampler and the target mouth as a sampling orifice. There have been many efforts since the 1970s to predict aspiration efficiency (*AE*, ratio of inhaled concentration to mainstream concentration, also referred to as inhalability), such as those of Ogden and Birkett [32], Armbruster and Breuer [33] and Vincent and Mark [34]. Many aspects of *AE* have been studied, using manikin experiments [35-38], theoretical models [13, 39] and CFD simulations [11, 40]. Great discrepancy exists among empirical equations. For example, the International Standards Organization (ISO) convention assumes a continuous decline of inhalability with the increase of aerosol diameter, while according to the American Conference of Governmental Industrial Hygienists (ACGIH) the aspiration efficiency levels off at approximately 0.5 [36]. Many equations were derived under specific experimental settings, thus failing to consider every potential factor. Note that the largest droplet diameters considered in the above-mentioned studies were 185 μm, which is close to the large droplet range as defined here. Although exhaled droplets can be as large as 1 mm, such sizes are rare, and these droplets are probably not as infectiously important as finer droplets, which contain most of the viruses.

The combined effect of mainstream air flow and sampling inhalation is that the streamlines first diverge when approaching the sampler, and then converge into the orifice. Dunnett and Ingham [41] established a 3D inhalation model with a spherical blunt sampler, which was shown in satisfactory agreement with the experimental results by Ogden and Birkett [32], as shown in Figure 4a. In contrast to the other models mentioned above, a complete set of influential factors was considered, without restrictions on the velocity and droplet size, thus providing important theoretical insights. Therefore, this inhalation model was adopted here.

**Figure 4.**
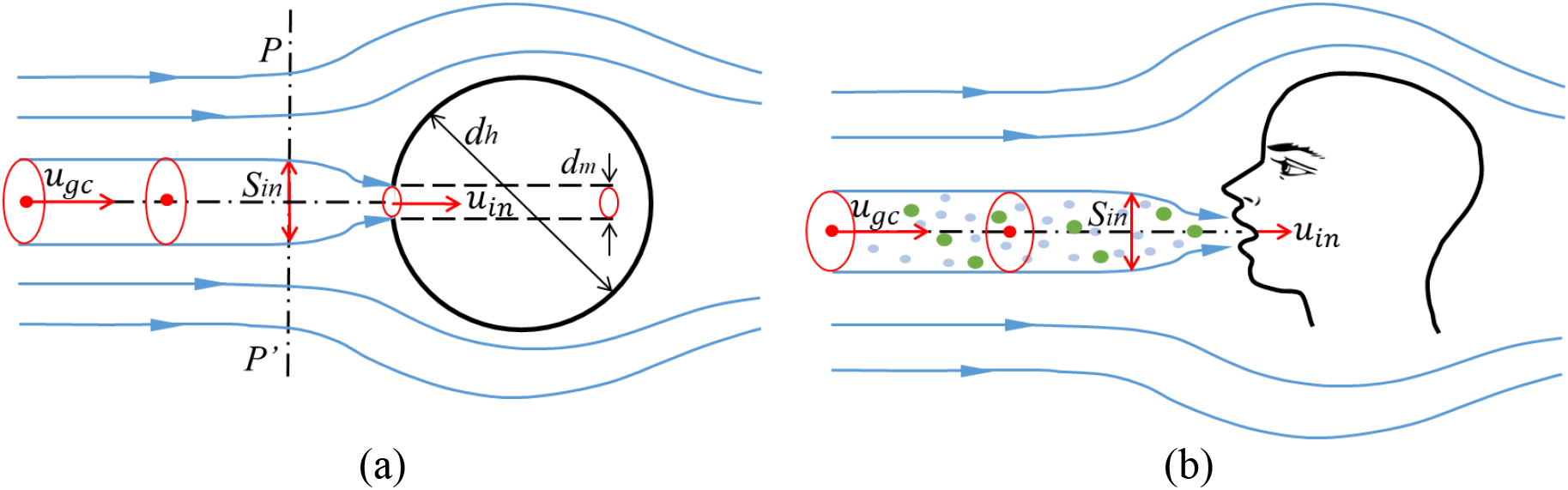
(a) Schematic diagram of aerosol sampling process with a spherical blunt sampler; (b) Illustration of the short-range airborne route with mouth inhalation. Note that only a fraction of the droplets entering the inhalation zone would eventually be inhaled.

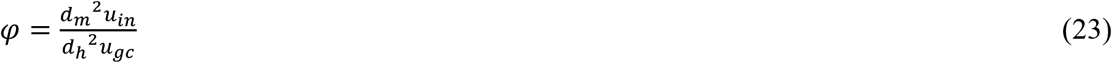

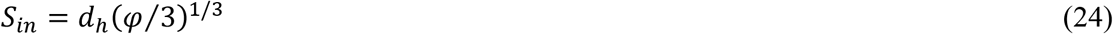

where φ is the sampling ratio for the axisymmetric flow system; *u*_*in*_ is the inhalation velocity (1 m/s); *S*_*in*_ is the width of the region on the sampler enclosed by the limiting stream surface. Note that we only consider the specific situation in which the negative mouth normal direction and the air flow direction are identical.

*IF* is simply the proportion of droplets that can enter the inhalation zone enclosed by the limiting streamlines (Figure 4b).

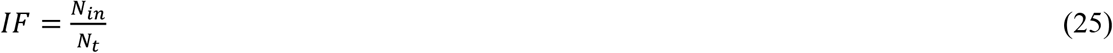

where *N*_*in*_ is the number of droplets entering the inhalation zone; *N*_*t*_ is the total number of released droplets at the mouth of the infected.

The inhalation zone is taken as a circular region in front of the target mouth with a diameter

*S*_*in*_ as calculated by the aspiration efficiency model (Equation (24)). We can obtain *N*_*in*_ by determining whether a particular droplet is within the inhalation zone. The position of the inhalation zone is also where the divergent centrelines become convergent (plane *PP’* in Figure 4a). We ignore the small gap between the susceptible person’s mouth and the *PP’* plane. A fraction of these *N*_*in*_ droplets will deposit on the target surface, while the others will be inhaled.

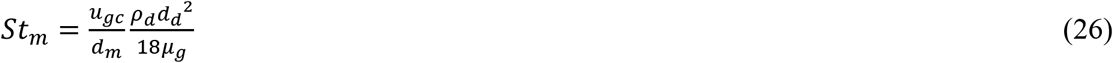

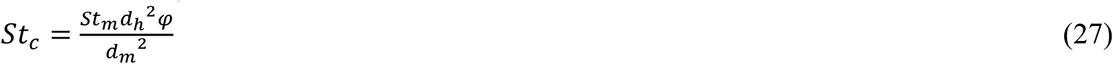

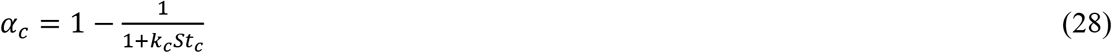

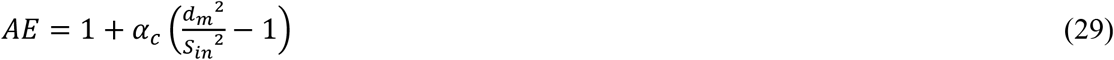

where *St*_*m*_ is the Stokes number for the mouth; *St*_*c*_ the Stokes number in the convergent part; *α*_*c*_ the impaction efficiency in the convergent part; and the constant *K*_*c*_ equals 0.3 when directly facing the incoming flow. Note that *u*_*gc*_ was adopted as the oncoming flow velocity for inhalation calculation, since the jet curvature within 2 m was negligible.

## 3. Results

### 3.1 Medium size droplets (75 to 400 μm) travel the shortest distance

Figure 5a shows the maximum travel distance for various droplet sizes. Note that the travel distance here was defined as the longest distance at which droplets could be detected, so the maximum value is perforce 2 m in this study, which does not necessarily mean that these droplets could not travel further. The shortest distance was travelled by droplets with diameters of approximately 112.5 to 225 μm for talking and 175 to 225 μm for coughing. In general, within the close range (2 m) studied, the small size group (<75 μm) would follow the air stream, being widely dispersed. The medium size group (75 to 400 μm) would be dominated by gravity, falling rapidly to the ground. The very large size group (>400 μm) would be dominated by inertia and travel a longer distance. The trend of our results is consistent with the CFD results by Zhu et al. [42] and Sun and Ji [43], although they did not quantify it. In the above discussion of travel *distance*, we noted the effect of size groups to avoid confusion with the relationship between droplet size and *exposure* in later discussion.

**Figure 5.**
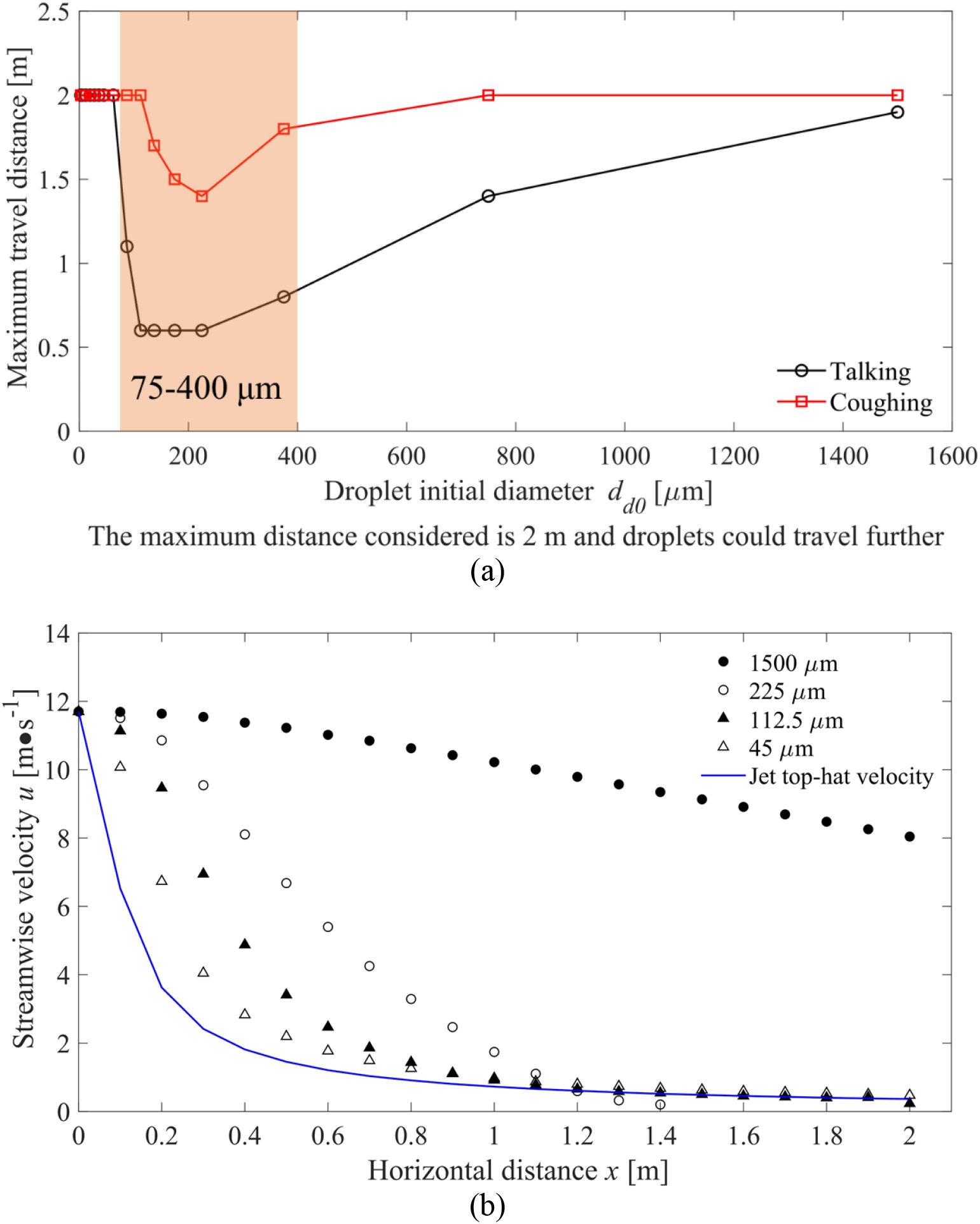
(a) Predicted maximum travel distances for various sizes of droplets during talking and coughing activities. Note that we consider a maximum travel distance of 2 m. (b) Differences between the averaged streamwise velocity of droplets with diameters of 1500, 225, 112.5 and 45 μm after being released, and the jet velocity based on top-hat profile at various distances from the mouth of the infected during coughing.

To elucidate the above results, the calculated velocities of air and droplets in a cough jet are compared for four droplet sizes: 1500, 225, 112.5 and 45 μm (Figure 5b). The smaller droplets (45 μm) have a very rapid momentum-response time (Table 1), which allows them to quickly follow the exhaled air stream, whilst the larger droplets (1500 μm) maintain their own velocity due to their more sluggish momentum-response time. This suggests that over a short distance, very large droplets are unlikely to settle.

**Table 1.**
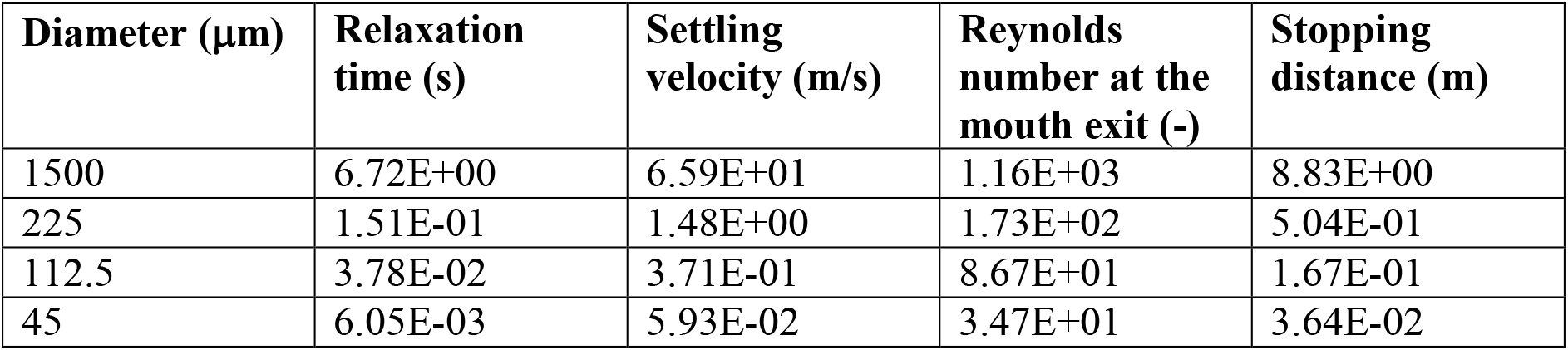
Droplet dynamics comparison in a cough jet.

| Diameter ($\mu\text{m}$) | Relaxation time (s) | Settling velocity (m/s) | Reynolds number at the mouth exit (-) | Stopping distance (m) |
| --- | --- | --- | --- | --- |
| 1500 | 6.72E+00 | 6.59E+01 | 1.16E+03 | 8.83E+00 |
| 225 | 1.51E-01 | 1.48E+00 | 1.73E+02 | 5.04E-01 |
| 112.5 | 3.78E-02 | 3.71E-01 | 8.67E+01 | 1.67E-01 |
| 45 | 6.05E-03 | 5.93E-02 | 3.47E+01 | 3.64E-02 |

### 3.2 Significant impact of exhalation velocity on travel distance and size change

Evaporation and falling processes compete after droplets are expelled from the mouth, so a critical size exists at which the falling time equals the evaporation time [44]. Various ambient environments (i.e., RH, temperature, etc.) and initial injection velocities also influence the droplet thermodynamics [45]. In these two studies by Wells [44] and Xie et al. [45], droplets were assumed to be perfect spheres that evaporated to a final diameter because of the existence of insoluble solids [20]. The change in dimensionless droplet diameter was compared for several typical initial sizes covering the whole range studied (see Figure 6).

**Figure 6.**
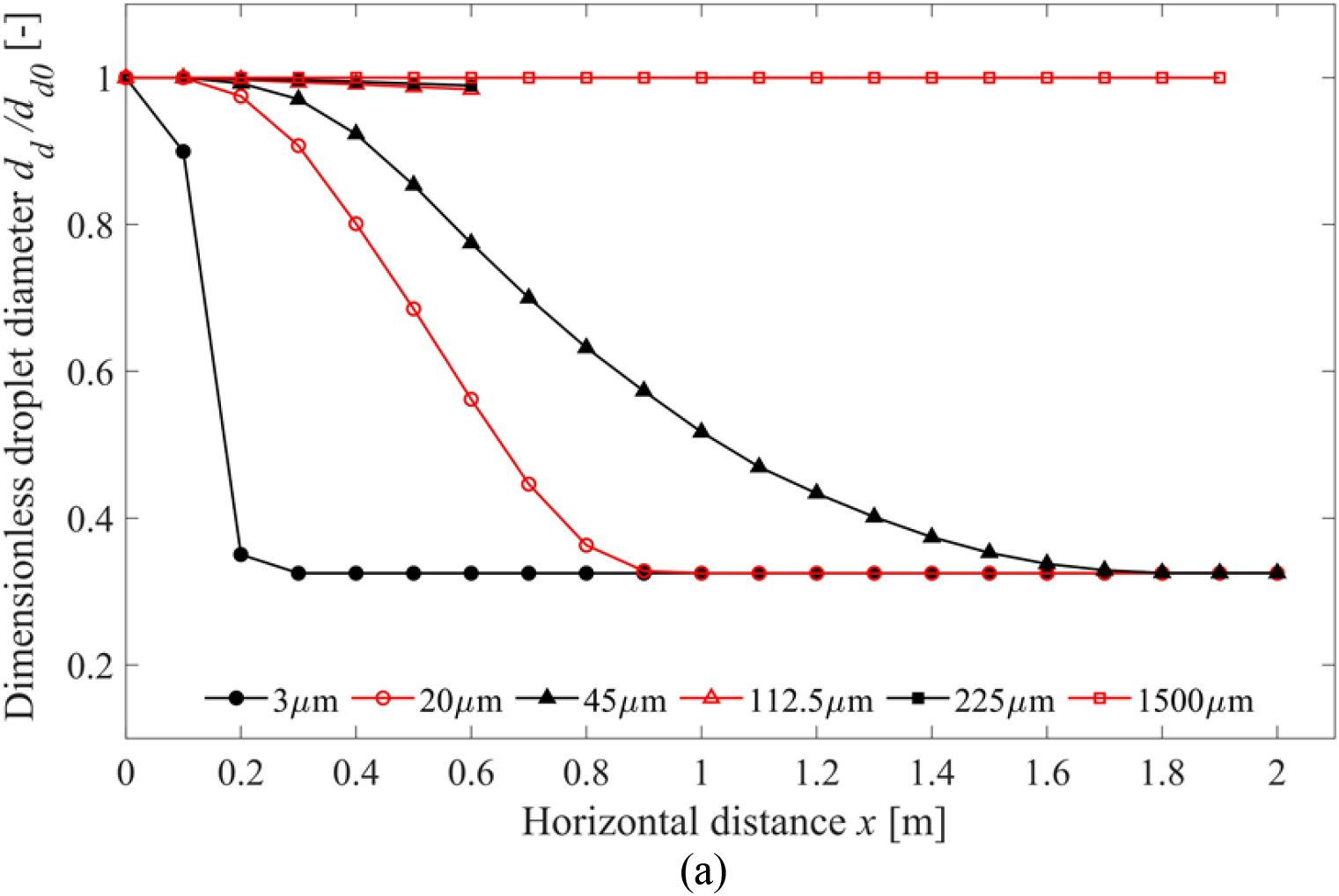
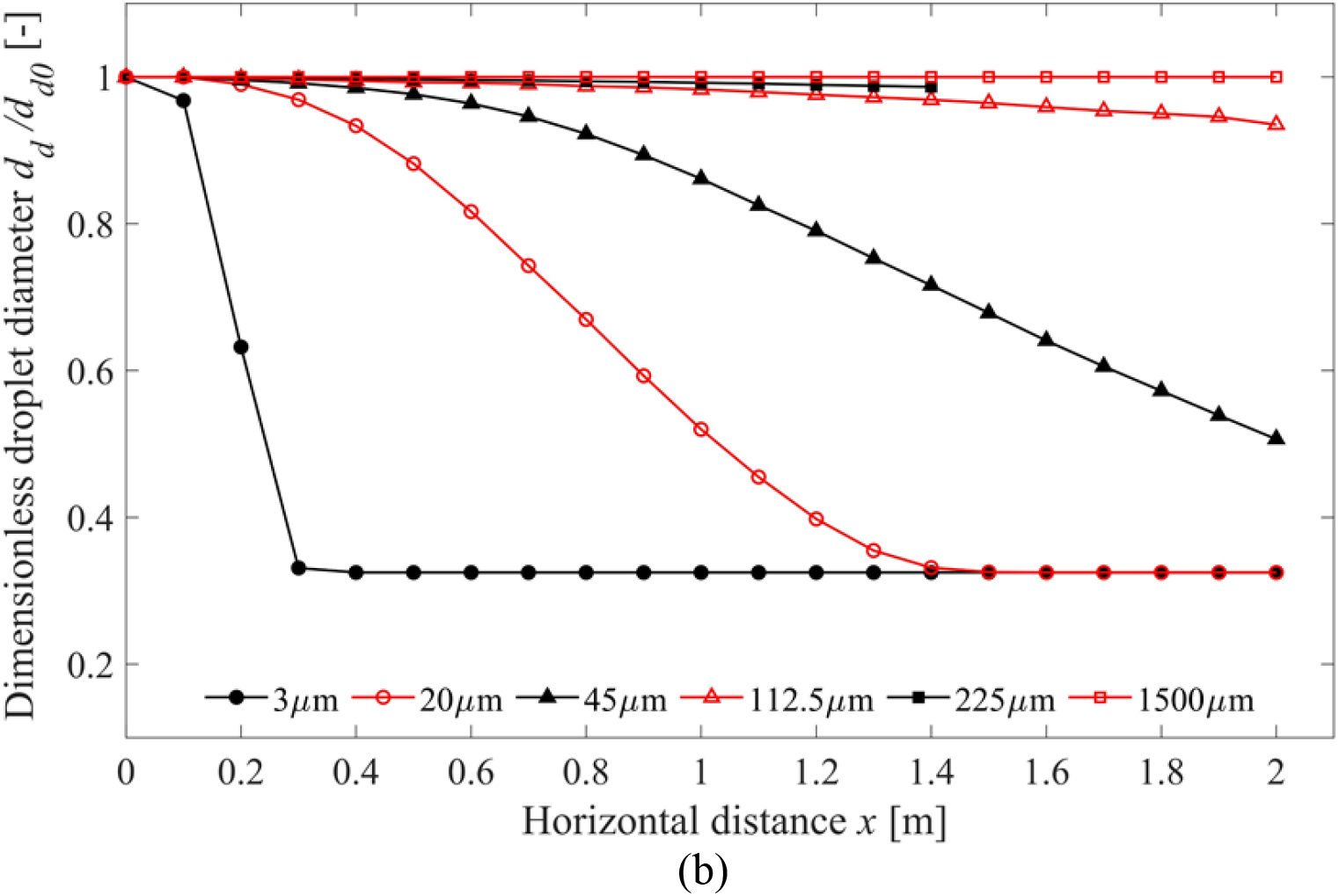
Changes in dimensionless droplet diameter while travelling away from the mouth of the infected for (a) talking; (b) coughing. Note that once all simulated droplets of a particular size land on the ground, no size is shown.

Because it was assumed that all droplets shared the same initial solid volume ratio, the final dimensionless diameter value remained constant for each size. For the assumed droplet composition here, the final size is 32.5% of the original diameter. Exhalation velocity was shown to have a significant impact on droplet travel distance for the medium size group (75 to 400 μm). The droplets of 112.5 μm and 225 μm in diameter travelled more than twice as far due to coughing than due to talking. Although the medium and very large droplets continued to shrink throughout their 2-m flight, the small droplets evaporated much more quickly, reaching their final size at some distance short of 2 m. The 3-μm droplets shrank rapidly within the first 0.1 m.

### 3.3 Total exposure

The total exposure of the susceptible person is shown in Figure 7 as a function of distance from the infected. To facilitate comparison, the exposure profile drawn on a logarithmic scale is also included. As expected, the exposure generally decreases as distance increases for both the large droplet and short-range airborne sub-routes. As shown in Figure B4(a), the coughing inhalation zone is smaller than target mouth at 0.1-0.3 m. It is too soon for droplets to disperse widely within 0.3 m, so more of them would be encompassed into the inhalation zone with an increase of size. The short-range inhalation exposure increases from 0.1-0.3 m is due to the enlargement of inhalation zone area, which directly influences the inhalation fraction (*IF*). From 0.3 m on, the overall decrease of exposure is dominated by jet dilution.

**Figure 7.**
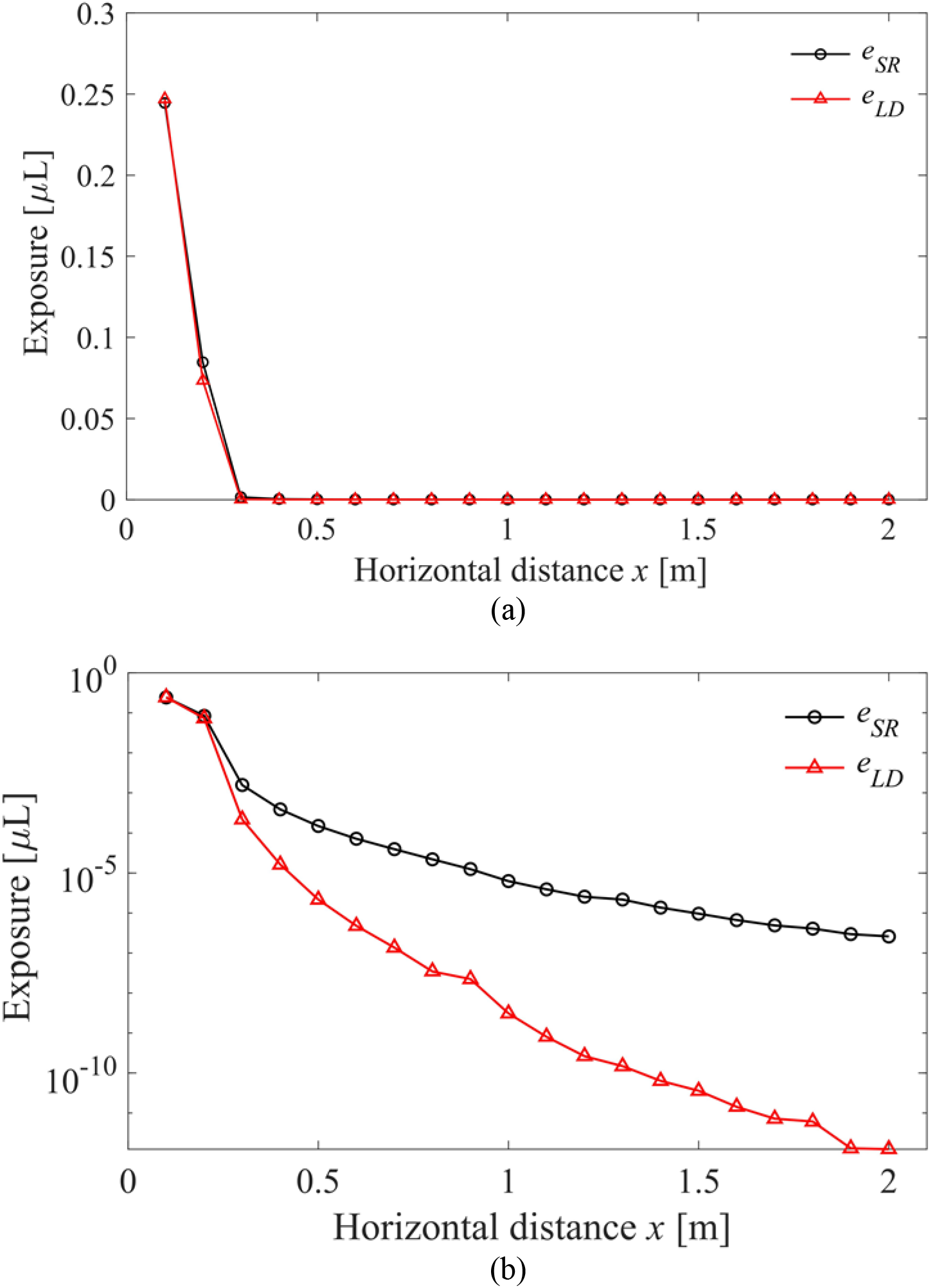
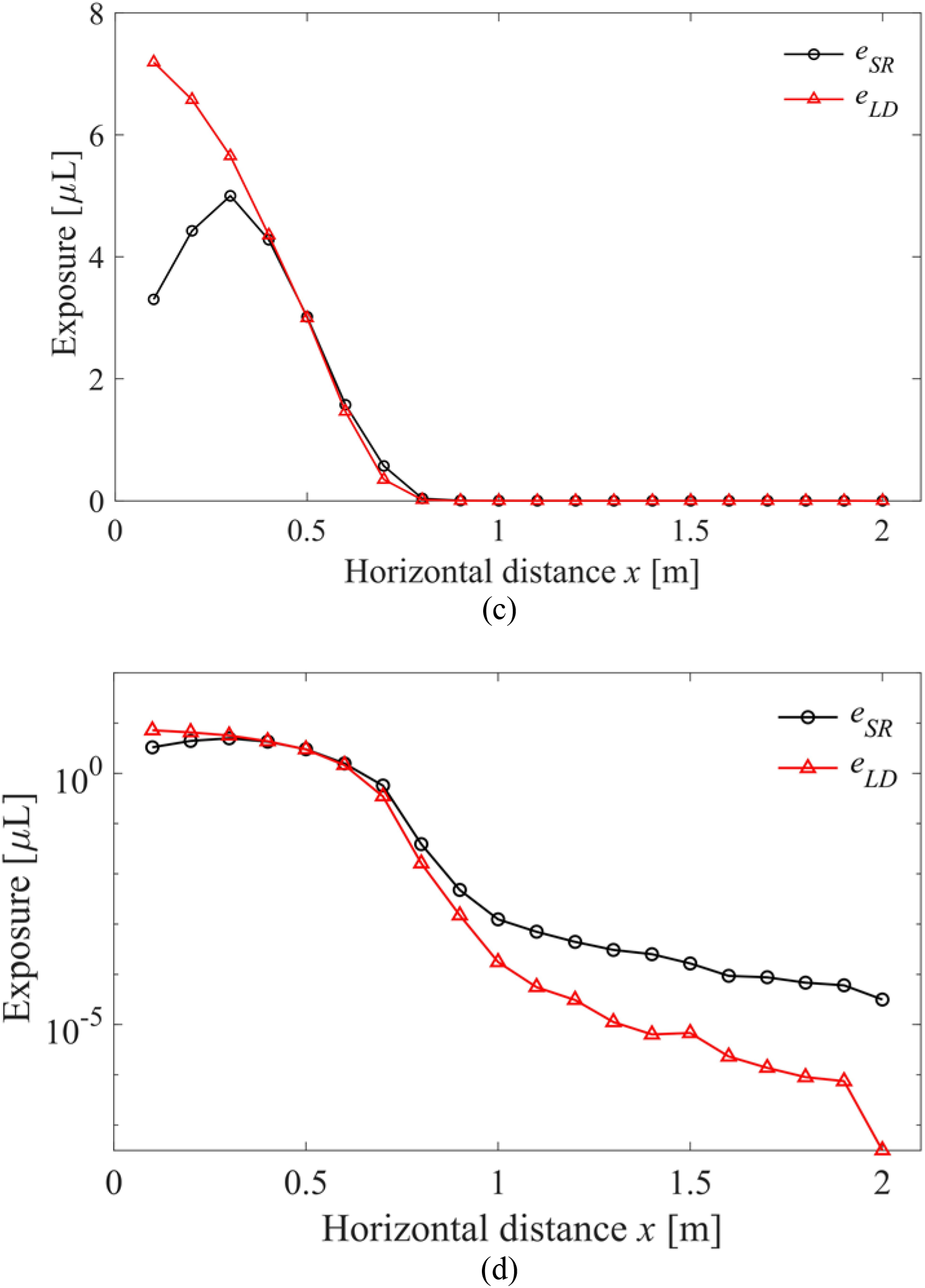
Total exposure for (a) talking (i.e. prolonged counting from ‘1’ to ‘100’) on normal scale; (b) talking (i.e. prolonged counting from ‘1’ to ‘100’) on logarithmic scale; (c) coughing once on normal scale; (d) coughing once on logarithmic scale.

As a whole, the exposure due to talking is an order of magnitude lower than that due to coughing for the situation considered here. The talking exposure was estimated based on prolonged loud speaking in which subjects were asked to count from ‘1’ to ‘100’, whilst coughing exposure was based on a single cough with the mouth initially closed. Given the same time period as for talking, coughing still causes a higher infection risk than talking considering coughing frequency of patients [46]. The total exposure value decreased by several orders of magnitude to almost zero at 0.3 m for talking and 0.8 m for coughing. A steep decline could also be detected in the logarithmic plots at the same distance. Notably, and unexpectedly, the short-range airborne route posed a greater exposure risk than the large droplet route, for both respiratory activities, at most distances in this close-range study, especially the longer distances.

### 3.4 LS exposure ratio

An *LS* ratio greater than unity (1) reveals a more significant role of the large droplet route. To better understand the influences of different droplet sizes, we subdivided the initial droplet size range into three segments for analysing *LS* exposure ratio: fine droplets smaller than 50 μm, intermediate droplets between 50 and 100 μm and large droplets greater than 100 μm.

Note that this classification differs from what we defined earlier (small <75 μm, medium 75-400 μm, very large >400 μm) in the analysis of travel distance. The *LS* ratio is shown as a function of distance in Figure 8. For the large droplet group, the exposure risk by the large droplet and/or short-range airborne routes dropped to zero beyond 0.5 m and 1.5 m for talking and coughing, respectively. Therefore, in Figure 8c, the data do not span the entire distance. The last two plots (c) and (d) are nearly identical, indicating that the large droplets dominate the overall exposure. This is to be expected because the droplet volume is proportional to the cube of droplet diameter, and thus the volume of a 750 μm droplet is 1.56 × 10^7^ times that of a 3 μm droplet. Figures 8a-c show that for larger droplets, the short-range airborne route becomes less important, as the *LS* ratio increases with droplet size. The *LS* ratio exhibited a quasi-exponential decay for droplets smaller than 100 μm, whilst for large droplets the ratio showed more fluctuation. A plateau from 0.4-0.6 m was notable. In this range, the inhalation zone diameter begins to experience a slower growth rate (Figure B4). For large droplets in Figure 8c-d, the averaged vertical coordinate is still within mouth; nevertheless, from 0.6 m on, they began to fall out of it. The fluctuation of the *LS* ratio for large droplets may also be due to the uneven initial droplet-size distribution in this range, as illustrated in Figure 2.

**Figure 8.**
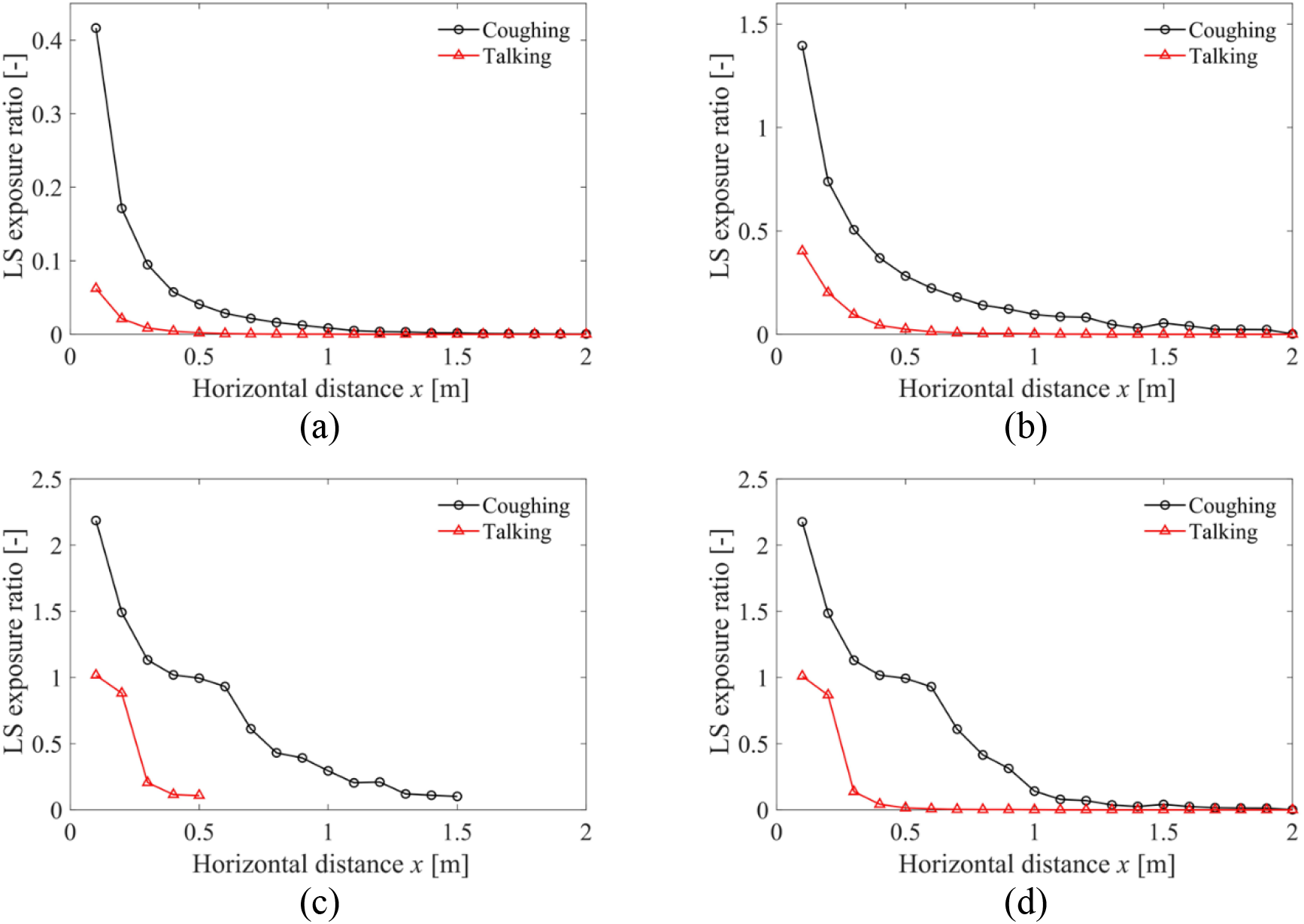
*LS* ratio for (a) <50 μm; (b) 50-100 μm; (c) >100 μm (0.1-0.5 m for talking and 0.1-1.5 m for coughing); (d) all sizes of droplets. Note: different vertical axis ranges are used.

The results obtained for the whole droplet size range in Figure 8d are interesting. We can conclude that the large droplet route is only dominant for talking within 0.2 m and for coughing within 0.5 m. The short-range airborne is much more important at the remainder of the close ranges studied here.

## 4. Discussion

### 4.1 The short-range airborne sub-route dominates the close contact transmission

Our calculation shows that in contradiction to what is commonly believed, intermediate and large droplets (including categories: 50 to 100 μm and >100 μm) are much less likely to be deposited on the lip/eye/nostril mucosa of a susceptible person than to be inhaled, unless the two are in very close contact. For the ideal situation that we have considered, the sphere within which large droplets dominate deposition is 0.2 m for talking and approximately 0.5 m for coughing. In all other situations, the short-range airborne route dominates exposure. The inhalation area is much larger for talking than coughing, which explains why the talking-induced short-range airborne route is more important than that for coughing (Figure B4). The difference in inhalation zone areas directly affects the membrane/inhalation ratio.

Reviewing the literature on large droplet transmission, one can find no direct evidence for large droplets as the route of transmission of any disease. It is known that the infection risk of many respiratory infections becomes higher when people come into closer contact. Flügge [6] pioneered the concept of large droplet transmission. He found that expiratory droplets contained bacteria and could not travel more than 1 or 2 m. Flügge [6] concluded that the expired droplets ‘*settled out in short distances and in brief time intervals, airborne infection seemed almost eliminated*’ [47]. The large droplet route became widely accepted after Chapin [7] developed his theory of the dominant contact transmission. Atkinson and Wein [24] suggested that large droplet transmission is less likely than formerly believed because close and unprotected exposure to direct expired air streams is rare. Our analysis disagrees with this point of view, instead showing that the insignificant role of large droplet transmission is due to the low rate of deposition even when direct expired air streams do exist.

It seems that we are the first to consider the dependence of the deposition behaviour on the Stokes number and that of the inhalation probability on the aspiration efficiency. Although these are important physical parameters of close contact exposure, they were not considered in previous studies.

Our work clearly shows that exposure due to the short-range airborne route dominates the overall exposure risk for droplets smaller than 50 μm. Note that our calculation of exposure is based on droplet volumes only. In directly comparing the two exposures for the purpose of discussing infection risk, we implicitly assume that the virus concentrations are the same in all sizes of droplets, which is unlikely. Indeed, one common supporting argument for large droplet transmission is that large droplets contain most of the infectious viruses, whilst fine droplets do not. This was recently found to be untrue: instead, studies have shown that smaller droplets have higher virus concentrations than larger droplets [48-49]. Zhou et al. [50], in experiments on captive ferrets, found that droplets less than 1 μm were not infectious, whilst those from 2 to 6 μm did transmit infection; larger droplets were not identified. The droplet sizes (after evaporation) considered in those studies were all very small. The most relevant droplet size range in this study is thus 0 to 50 μm (Figure 8a). In this range, the exposure due to the short-range airborne sub-route would be more than 2 times that due to large droplets even at a close distance of 0.1 m for coughing. For a typical inter-personal distance of 0.7 m [3], the same ratio for coughing becomes over 45. Note that we only compared the two sub-routes for talking and coughing separately, without considering the relative frequency of these respiratory activities. Face-to-face coughing is a rare event [24].

There is a need to test the variability in the concentration of viable viruses in expired air streams. For this purpose, new, more efficient samplers that can better preserve virus activity are necessary [48, 51].

### 4.2 Threshold droplet size for large droplet is not 5 or 10 μm, but 50-100 μm

Our calculation of the deposition efficiency clearly shows that droplets smaller than 100 μm are less likely to be deposited on the facial parts of the susceptible person (Figure 8), although it is not the main purpose of this paper to calculate the large droplet threshold size. However, this is an important concept that is relevant to our discussion of the dominant sub-route. In Figure 8, droplets at the point of release (i.e. mouth) are divided into three ranges: fine droplets (0-50 μm), intermediate sizes (50-100 μm) and large droplets (>100 μm). For the size range 0 to 50 μm, the droplets will be airborne in the expired air streams for the time scales that we consider here, particularly after evaporation.

Our calculation confirms that the size-dependent difference in the deposition efficiency of droplets on the face is one of the major reasons for the calculated differences between the two exposure routes. Droplets in the small size group (<75 μm), which can closely follow the air stream, have relatively low Stokes numbers and are unlikely to be deposited. The medium size group (75-400 μm) would land on the ground the soonest. Droplets in the very large size group (>400 μm) have the greatest potential for facial deposition and travel the greatest distance before falling to the ground.

Thus, the commonly assumed threshold droplet size of 5 or 10 μm is not only wrong, but intrinsically misleading. This assumption leads to the false conclusion that droplet transmission only applies to droplets larger than 5 μm. Our literature review shows that it was probably Garner et al. [52] who first suggested this droplet transmission lower boundary of 5 μm, without citing any reference. The WHO 2014 guideline [53] still defines droplets as ‘respiratory aerosols > 5 μm in diameter’. Siegel et al. [54] recognised that ‘observations of particle dynamics have demonstrated that a range of droplet sizes, including those with diameters of 30 µm or greater, can remain suspended in the air’. We distinguished the two sub-routes known as “large droplet” and “short-range airborne” according to the way the susceptible was exposed to (i.e., deposition and inhalation) in this study, and our determined size range also differs from the traditional droplet size range. Traditional term such as large droplet transmission may be misleading. However, more effort would be necessary for recognizing the threshold droplet size, and the precise transmission route(s) need to be reconsidered as more data become available.

### 4.3 Assumption of the dominant large droplet sub-route may hinder development and acceptance of alternative interventions

The effectiveness of surgical masks depends on the dominance of large-droplet transmission by droplets greater than 50 μm in diameter. A number of studies have questioned their effectiveness against influenza. Milton et al. [48] found that surgical masks could reduce viral copy numbers by 25-fold for droplets larger than 5 μm but only 2.8-fold for fine droplets smaller than 5 μm. The use of facemasks itself is not detrimental, but reflects a strong belief in the dominant role of large droplet transmission, due to which other possible interventions are likely to be neglected.

Mechanistically, the use of surgical masks by an infected can ‘block’ or ‘kill’ expiratory jets; that is, the expired air is initially blocked within the facial cavity of the mask of the infected before eventually leaking out to the environment through the mask itself or the gaps on either side. The momentum of the blocked expired jet becomes so weak that it is most likely to be captured by the body plume of the infected. The body plume carries the weakened expired stream into the upper level of the indoor space, which eventually becomes a part of the room air, contributing to the long-range airborne route, which is expected to be much weaker than the short-range airborne route.

Importantly, the expired air streams have a velocity much greater than the typical indoor air flows (0.2 m/s), hence the room air flows do not significantly alter the expired jet trajectory. Hence, general ventilation cannot prevent transmission by the short-range airborne route [9].

Personalised ventilation systems may be effective here because they provide filtered and safe air directly to the breathing zone of the susceptible person [55-57]. Personalised ventilation devices can be installed at fixed places such as office chairs, desks or computers, enabling occupants to control its temperature, flow rate and direction [55]. However, for people without fixed workplace, no existing ventilation strategy is currently available for mitigating the short-range airborne route, and innovative new ideas are needed.

### 4.4 Difference between the short-range airborne and large droplet route

In this study, we considered the short-range airborne route and the large droplet route separately as two processes. The susceptible person was assumed to hold his or her breath with mouth open for the large droplet route and inhale orally for the short-range airborne route. The situation of coexistence of the two routes was also calculated, where an imaginary plane at the target mouth was responsible for the large droplet sub-route; see Appendix A for a summary of the important results.

It is important to uncover the mechanistic details of the difference between the airborne and large droplet routes. The fate of droplets after entering the human body through respiratory activities seems to depend on their size. Different droplet sizes lead to differences in deposition efficiency at different sites (i.e., head airways, tracheobronchial region or alveolar region) [58]. According to Carvalho et al. [59], particles between 1 and 5 μm are deposited deep in the lungs, whilst those larger than 10 μm are generally deposited in the oropharyngeal region, and particles smaller than 1 μm are exhaled. The response dose can also be region-sensitive for drug delivery [60] and potential hazard [61]. If we consider the final fate of infectious droplets, their destiny is deposition, whether in the head airways or in alveoli, via inertia impaction, sedimentation or diffusion. The relative probabilities of the short-range airborne route and the large droplet route may depend on processes external to the body, implying that disease prevention measures should focus on the ambient air streams.

We focused on the jet and droplet dynamics outside the human body in this study. The large droplet and short-range airborne routes become indistinguishable at the target mouth plane when considering both sub-routes simultaneously. As shown by Anthony and Flynn [11] using CFD, particles larger than 5 μm can be deposited on the inside surface of the lips due to gravity settling. If such a CFD approach is used, one may define inhalation more precisely by only including those particles that go through the area ‘between the lips’. Here we considered all particles that were ‘directed toward the mouth’ [11], which may be the upper bound of aspiration by inhalation. When a droplet passes through the mouth orifice, we cannot rigorously determine whether it is due to deposition or inhalation, which makes it meaningless to attempt to distinguish between them at the mouth plane. We therefore presented the results of the large droplet and short-range airborne routes as two separate processes in the main text. Note that when the two sub-routes co-exist (Appendix A), the major difference from the situations presented in the main text is that once a particle is inhaled, the particle is no longer available for deposition. The predicted range of dominance of the short-range airborne route was extended slightly to 0.3 m for talking and 0.9 m for coughing (Appendix A), although large droplet route becomes more important in the coexistence case. However, a more careful redefinition of the short-range airborne route and the large droplet route will require additional data.

### 4.5 Limitations of the study

Despite the valuable findings, our study still has the following limitations. First, exposure (μL) was used as the criterion of infection based on the assumption that every unit volume of droplet contains the same amount of activated viruses. Nevertheless, according to Lindsley et al. [62], most (∼65%) virus RNA was contained in droplets smaller than 4 μm expelled by coughing, which indicates a higher risk in the respiratory range.

Although the exclusion of droplets smaller than 3 μm would exert negligible influence on exposure given their small droplet volume, significant implications may exist when virus concentration variation is considered. The critical infective dose was also not considered. Future work could be done from a more biologically informed perspective based on the exposure results. Second, the number of simulated droplets was relatively small. Because *MF* and *IF* are statistical probability values, a larger number of droplets, if possible, would give more robust results. Third, the worst-case scenario of mouth inhalation and that of deposition were studied, which may deviate slightly from realistic situations. Such worst scenarios might occur during face to face conversations, but data on the frequency of its occurrence is not available. Although the effects associated with nose-versus-mouth breathing and facial structural features are weak [36], a more detailed nose inhalation model is still desirable. Our two nostrils are very close to each other, and they mostly face downward at a certain angle.

During the nasal inhalation, the configuration of the inhalation zone would be distorted by one another. Exposure due to both inhalation and deposition was estimated using existing empirical formulas assuming a spherical head shape. Other factors like relative subject height, face-to-face angle and mouth covering may greatly affect the exposure results.

Different indoor airflow patterns due to different air distribution strategies and human body thermal plumes, which can disperse droplets, would also cause discrepancies, especially at farther distances. Improved experiments and CFD simulations are needed to investigate the influence of potential factors under more realistic contexts.

Finally, only two transmission routes were considered in our work. Because the mucous membranes are small in area relative to the total frontal area of the head, most exhaled droplets are likely deposited on other regions like cheeks, neck or hair. These deposited droplets might be touched by the susceptible person’s own hands, which subsequently touch his or her mucosa, resulting in self-inoculation. Recent data by Zhang et al. [63] show that people touch their face very frequently. Facial deposition and touch may contribute another potential transmission route in close contact, which is worth exploring in future.

## 5. Conclusions

This is probably the first study in which the large droplet route, traditionally believed to be dominant, has been shown to be negligible compared with the short-range airborne route, at least for expired droplets smaller than 100 μm in size at the mouth of the infected. The exposure due to short-range airborne transmission surpasses that of the former route in most situations for both talking and coughing. The large droplet route only dominates when the droplets are larger than 100 μm, within 0.2 m for talking and 0.5 m for coughing. The smaller the exhaled droplets, the more important the short-range airborne route. The large droplet route contributes less than 10% of exposure when the droplets are less than 50 μm at a distance greater than 0.3 m, even for coughing. For the direct face-to-face configuration, exhaled air streams begin to cover the nostrils of the susceptible person from 0.2 to 0.3 m and the eyes from 0.4 to 0.5 m. While talking, more droplets are deposited on the eyes at long distances due to a larger jet trajectory curvature (Appendix B). Exposure decreases as the interpersonal distance increases for both large droplet and short-range airborne sub-routes.

Short-range airborne transmission is dominant beyond 0.2 m for talking and 0.5 m for coughing. Within the 2-m interpersonal distance, the shortest distance is travelled by droplets of approximately 112.5 to 225 μm in size for talking and 175 to 225 μm for coughing. The smaller droplets follow the indoor air stream, whilst the larger droplets are dominated by their inertia and travel a longer distance.

The work presented here poses a challenge to the traditional belief that large droplet infection is dominant. Because the short-range airborne route is dominant for both talking and coughing according to the results here, novel methods of personalised ventilation during close contact are worth considering as a strategy for disease control.

## Data Availability

Only mathematical models are presented and the original modelling data may be obtained by contacting the authors.

## Acknowledgements

This work was supported by a General Research Fund (grant number 17202719) and Collaborative Research Fund (grant number C7025-16G), both provided by the Research Grants Council of Hong Kong.

## Appendix A. Evaluation of LS ratio considering the coexistence of the two sub-routes

When considering the coexistence of the large droplet and short-range airborne routes, an imaginary plane was assumed at the target mouth. Droplets deposited on the plane were assigned to the large droplet route, and those filtering through it were assigned to short-range airborne transmission.

The short-range airborne exposure is still calculated as:

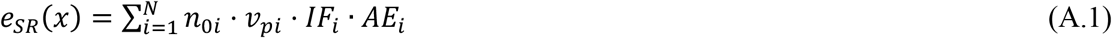

When the large droplet route and short-range airborne route co-exist, the droplet deposition behaviours are expected to be affected by inhalation flow. Based on whether droplets exist simultaneously both on facial membranes and in the inhalation zone, we divided the large droplet exposure into two parts, where the total large droplet exposure is the sum of them. *e*_*LD*1_(*x*) represents the case when droplets are outside the inhalation zone, whilst *e*_*LD*2_(*x*) indicates that facial mucous membranes overlap with inhalation zone. The membrane fraction (*MF*) and deposition efficiency (*DE*) also change accordingly.

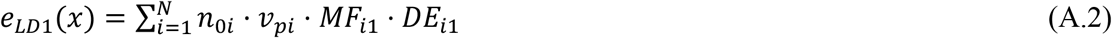

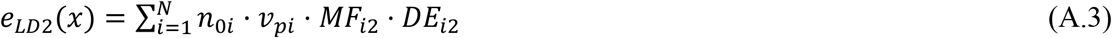

*DE*_*i*1_ remains the same as defined in Equation (21). Unlike the original inhalation model, when the short-range airborne and large droplet routes co-exist, an imaginary plane is included at the target mouth. Therefore, we made a small change to the original model, such that *DE*_*i*2_ equals α_c_, which is the impaction efficiency (Equation (28)). As *AE*_*i*_ and *DE*_*i*2_ affect each other in the convergent part of the air stream, *AE*_*i*_ equals 1 - *α*_*c*_ accordingly.

The results of the estimated total exposure and *LS* ratio are shown in Figure A1 and Figure A2 respectively.

**Figure A1.**
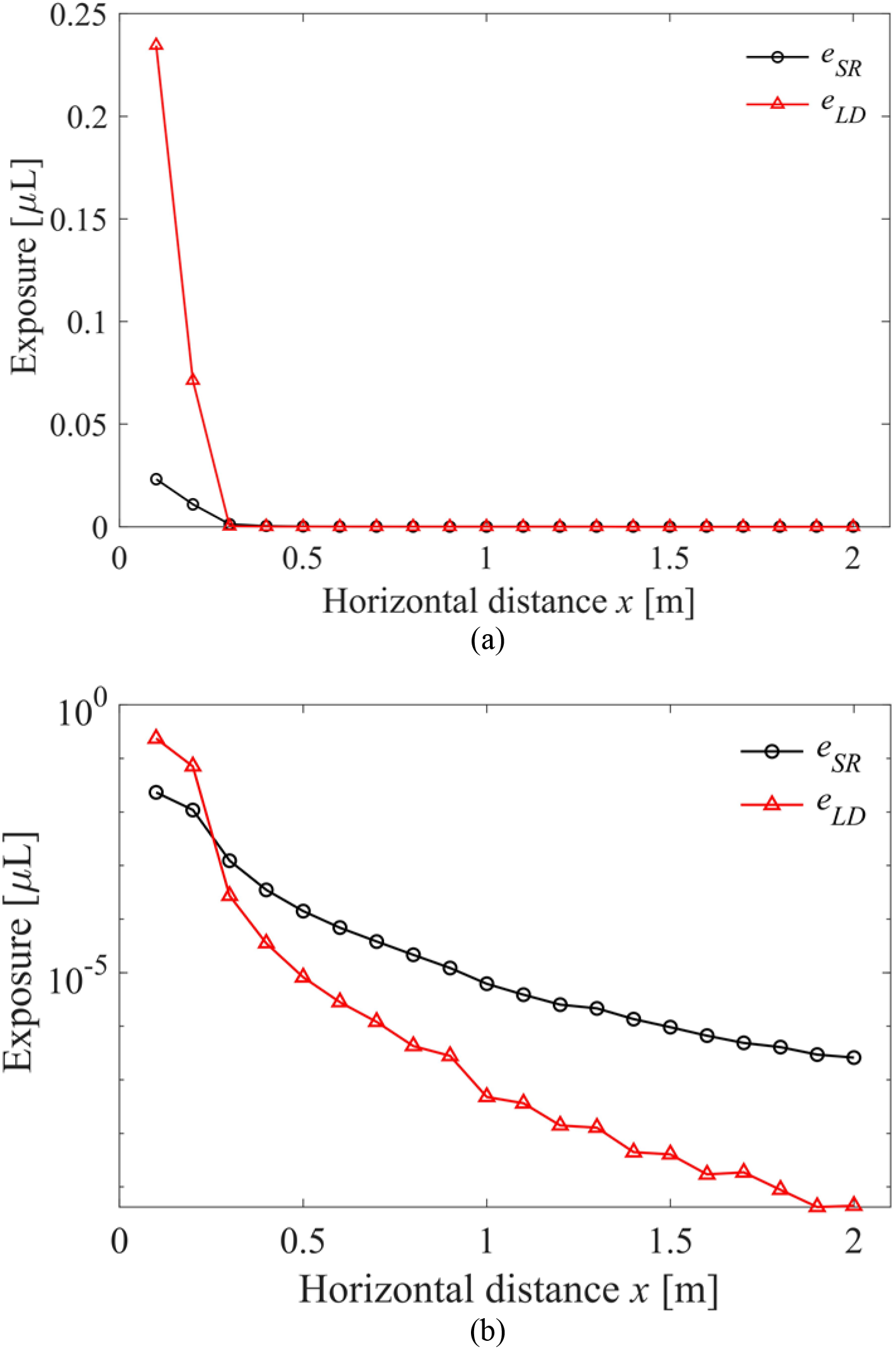
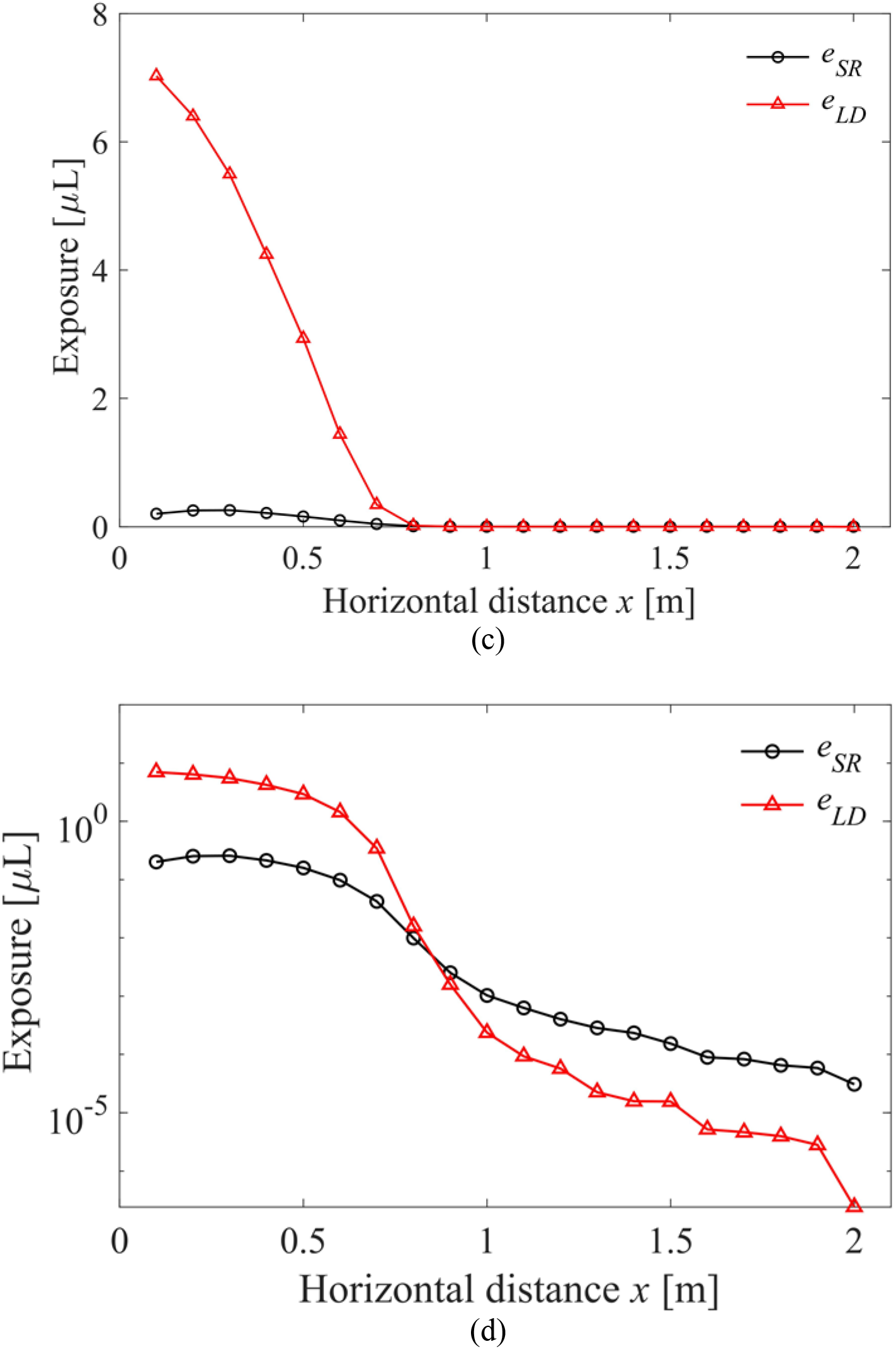
Total exposure for (a) talking (i.e. prolonged counting from ‘1’ to ‘100’ once) on normal scale; (b) talking (i.e. prolonged counting from ‘1’ to ‘100’ once) on logarithmic scale; (c) coughing once on normal scale; (d) coughing once on logarithmic scale.

**Figure A2.**
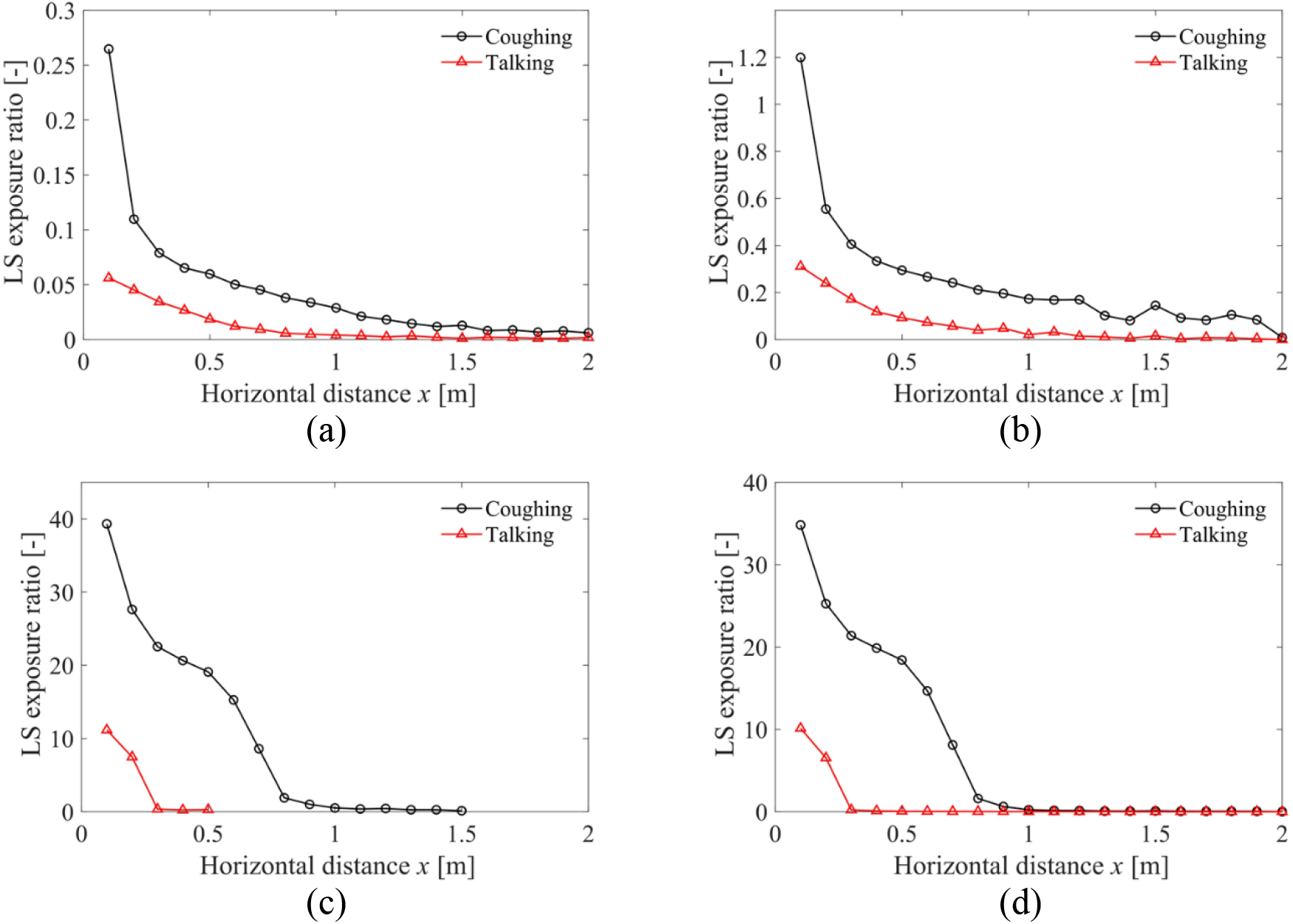
*LS* ratio for (a) <50 μm; (b) 50-100 μm; (c) >100 μm (0.1-0.5 m for talking and 0.1-1.5 m for coughing); (d) all sizes of droplets. Note different vertical axis ranges are used.

As a whole, the same trend was observed as in Figure 8, although short-range airborne sub-route becomes slightly more important for droplets smaller than 100 μm while large droplet route is dramatically more significant for those larger than 100 μm. The *LS* ratio values for talking/coughing all resemble each other for all droplet sizes, except that a slower decay was observed for coughing from 0.3 to 0.5 m. In this range, the inhalation zone diameter begins to experience a slower growth rate (Figure B4). For large droplets in Figure A2c-d, the averaged vertical coordinate is still within mouth; nevertheless, from 0.6 m on, they began to fall out of it. The fluctuation of the *LS* ratio for large droplets may also be due to the uneven initial droplet-size distribution in this range as illustrated in Figure 2.

## Appendix B. Deposition and aspiration

Statistically, our defined membrane fraction (*MF*) and inhalation fraction (*IF*) are case-sensitive probabilities smaller than 1. The values differ with the relative height of the target and source, face features, head direction and inhalation velocity. As mentioned above, the worst-case scenario was considered in this study. For the current specific case, *MF* and *IF* varied with distance and droplet size as demonstrated in Figure B1. Note that different legends are used. *MF* and *IF* dropped to approximately zero for large droplets at long distance. Figure B1c and d show that the talking *IF* and coughing *IF* differ considerably at close range (<0.5 m). Although the values for talking were dispersed uniformly across the whole size range, the maximum values appeared for large droplets, as highlighted at the left top corner. The overall trend of *MF* resembles that of *IF* for both talking and coughing. This indicates that higher exhalation velocities would affect the large droplet behaviours, which in turn influences exposure. A clear boundary can be detected for both talking *IF* and talking *MF*, where medium and large droplets begin to fall out of the jet region with a sharp decrease of their vertical coordinates. The critical size was around 62.5 μm.

The ratio of inhaled/deposited droplets for talking and coughing as a function of distance is shown in Figure B2. Inhaled droplets were one order of magnitude more numerous than deposited droplets, and exposure to inhaled droplets was greater for coughing. For both talking and coughing, the inhaled droplet number followed the same distribution pattern as the exhaled droplet number shown in Figure 2; the peak value appeared at a smaller droplet diameter of 12 μm. In contrast, the trend of droplet deposition was totally different.

Compared with inhalation, deposition is more distance-determined, with the deposited droplet number dropping to almost negligible beyond 0.3 m for talking and 0.8 m for coughing.

Because larger droplets have a larger Stokes number, it becomes easier for them to be deposited on the human face. Thus, the number of deposited droplets aggregated at the medium-large size range. Compared with talking, for coughing the deposition fraction showed a much slower decay with distance.

It is also worth investigating exactly where the droplets fall. We compare the deposition number percentage of each facial membrane for 3 μm and 36 μm droplets in Figure B3. Exhaled droplets began to cover nostrils from 0.2 to 0.3 m and the eyes from 0.4 to 0.5 m. The mouth became less important as the distance increased. Because of the lower exhalation velocity, the trajectory of the jet curved upward more obviously for talking than for coughing. Therefore, more droplets deposited onto the eyes at longer distance due to talking. Because eye protection has been proven to reduce infection via the ocular route, the use of masks with goggles or a face shield may be a promising policy.

**Figure B1.**
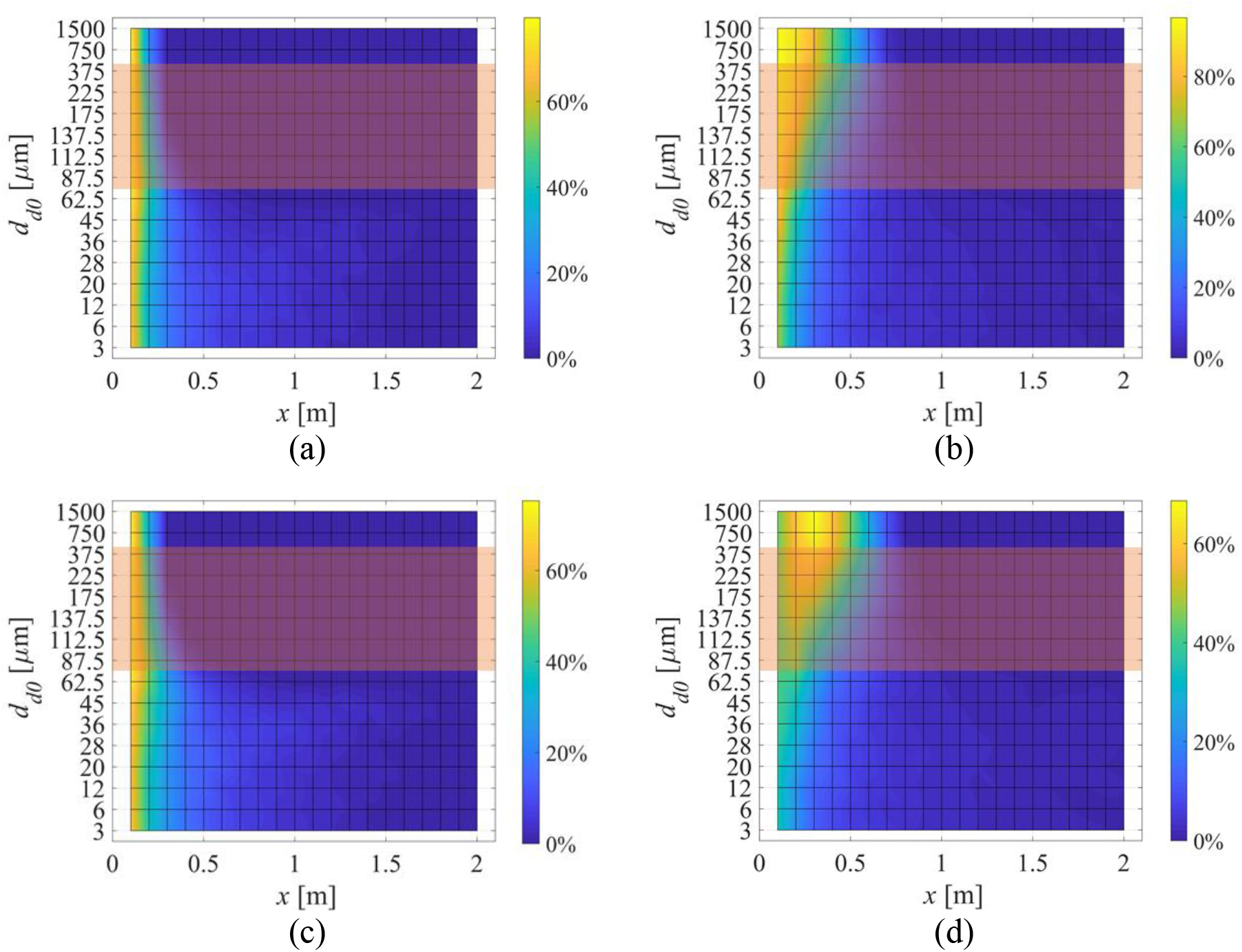
The calculated membrane fraction (*MF*) and inhalation fraction (*IF*) as a function of horizontal distance *x* and droplet initial size *d*_*d0*_. (a) Talking *MF*; (b) Coughing *MF*; (c) Talking *IF*; (d) Coughing *IF*.

**Figure B2.**
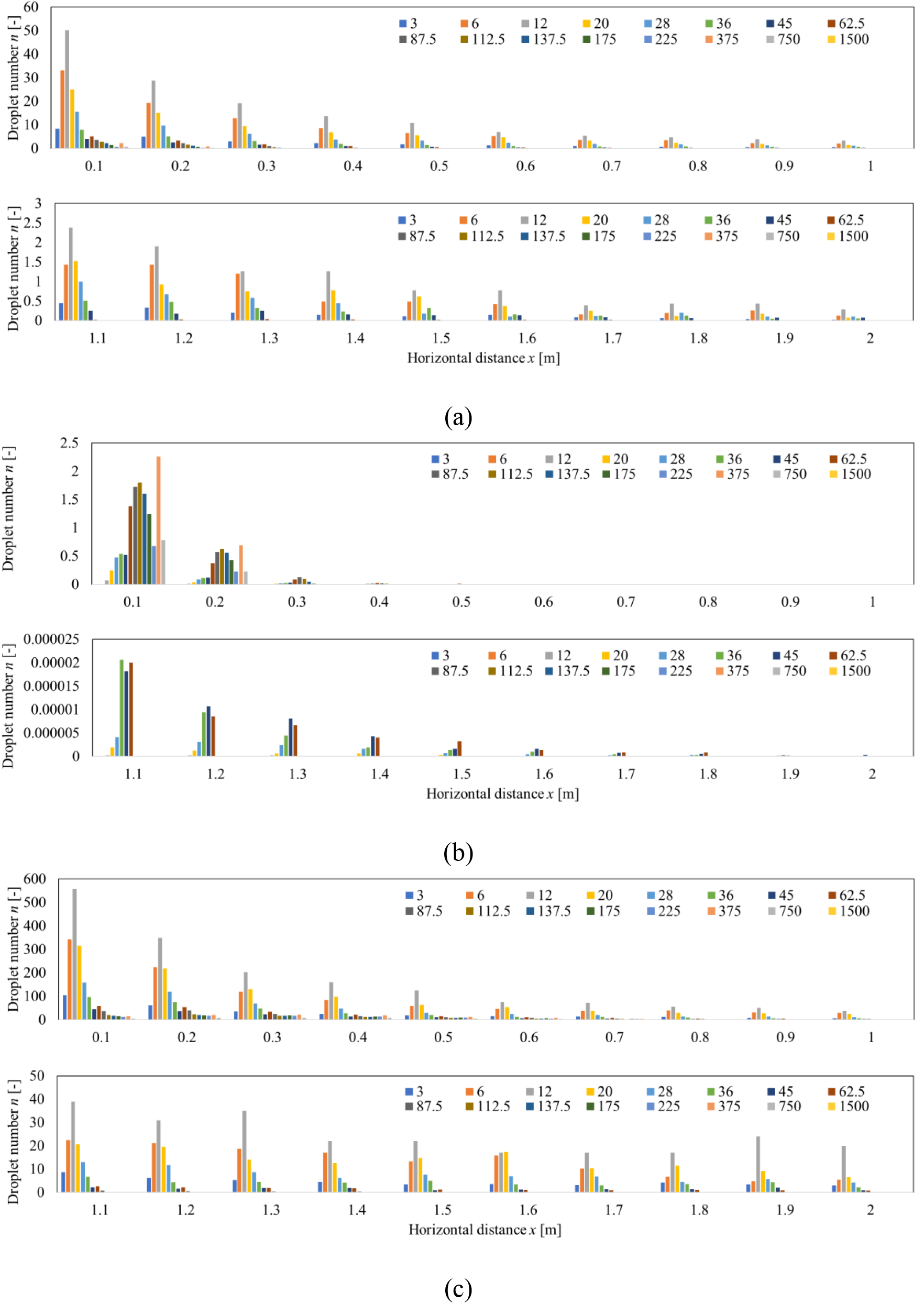
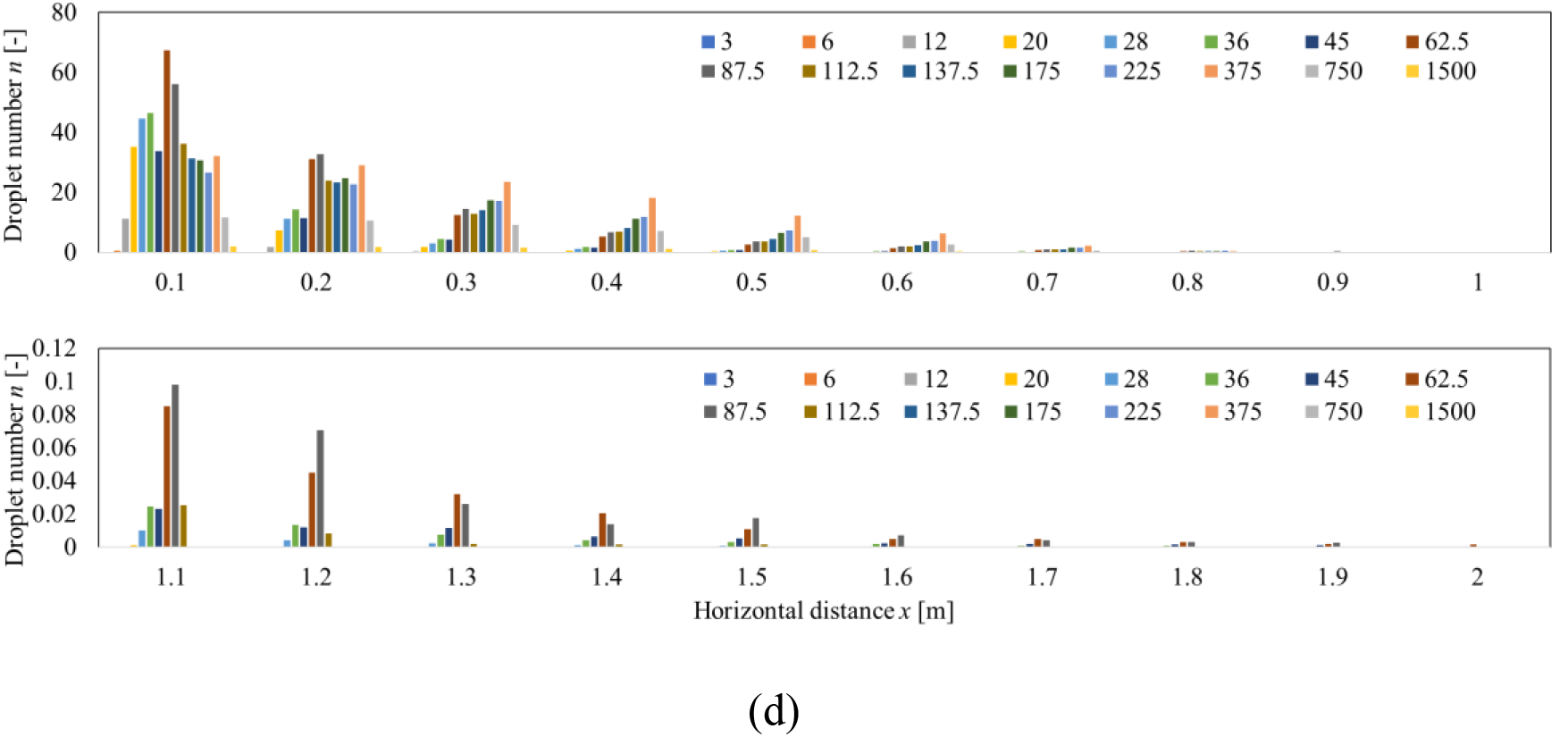
Number of inhaled/deposited droplets for talking by (a) Inhalation; (b) deposition on facial mucous membranes; and those for coughing by (c) Inhalation; (d) deposition on facial mucous membranes.

**Figure B3.**
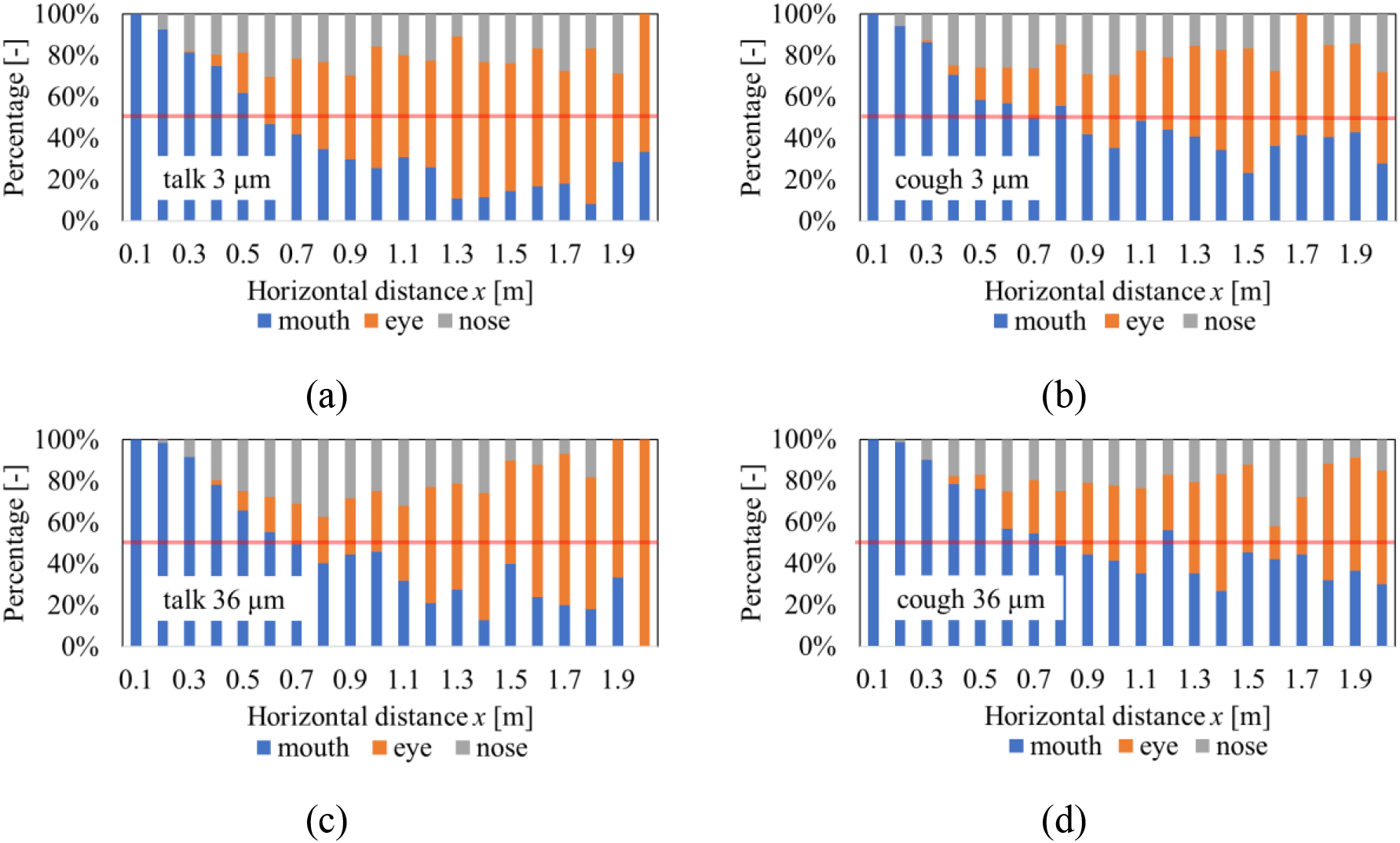
Percentage of droplet deposition location (a) 3 μm droplets for talking; (b) 3 μm droplets for coughing; (c) 36 μm droplets for talking; (d) 36 μm droplets for coughing.

**Figure B4.**
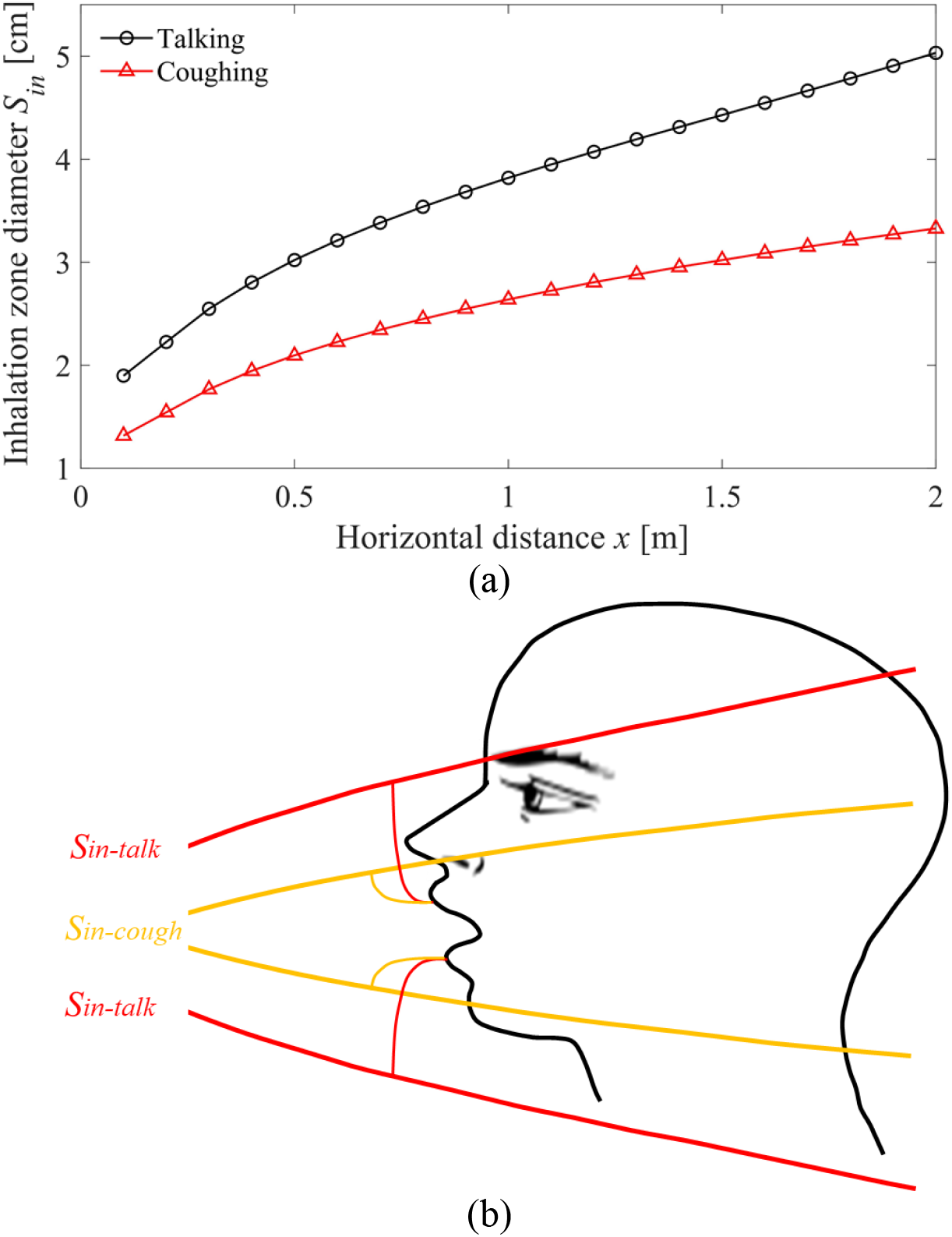
(a) Predicted diameter of the inhalation zone; see Figure 4b. (b) Illustration of inhalation zone diameter at 1 m relative to the possible location of the mouth opening of the susceptible person.

